# COVID-19 vaccination acceptance among healthcare workers in Germany

**DOI:** 10.1101/2021.04.20.21255794

**Authors:** Christopher Holzmann-Littig, Matthias Christoph Braunisch, Peter Kranke, Maria Popp, Christian Seeber, Falk Fichtner, Bianca Littig, Javier Carbajo-Lozoya, Christine Allwang, Tamara Frank, Joerg Johannes Meerpohl, Bernhard Haller, Christoph Schmaderer, on behalf of the CEOsys consortium

**Author notes:** **Correspondence to:** Christopher Holzmann-Littig and Christoph Schmaderer. Both first authors contributed equally to this work (Christopher Holzmann-Littig, Matthias Christoph Braunisch). Both last authors contributed equally to this work (Bernhard Haller, Christoph Schmaderer).

## Abstract

**Background:** Vaccination hesitancy is a serious threat to achieve herd immunity in a global and rapidly changing pandemic situation. Health care workers play a key role in the treatment of patients with Coronavirus disease 2019 (COVID-19) and in promoting vaccination in the general population. The aim of the study was to provide data on COVID-19 vaccination acceptance and barriers among healthcare workers in Germany to support health policymakers choosing specific vaccination campaign strategies.

**Methods:** An online survey was conducted among health care workers in Germany in February 2021. The survey included 55 items on demographics, previous vaccination behavior, trust in vaccines, physicians, pharma industry, and health politics as well as fear of adverse effects, assumptions on disease consequences, knowledge about vaccines, information seeking behavior and a short COVID-19 vaccine knowledge test.

**Results:** A total of 4500 surveys could be analyzed. The overall vaccination acceptance was 91.7%. The age group ≤20 years showed the lowest vaccination acceptance of all age groups. Regarding professional groups, residents showed the highest vaccination acceptance. Main factors for vaccination hesitancy were lack of trust in authorities and pharmaceutical companies. Personal and professional environment influenced the attitude towards a vaccination too. Participants with vaccination hesitancy were more likely to obtain information about COVID-19 vaccines via messenger services or online video platforms and underperformed in the knowledge test.

**Conclusions:** In conclusion, we found a high acceptance rate amongst German health care workers. Furthermore, several factors associated with vaccination hesitancy were identified which could be targeted in vaccination campaigns.

## Introduction

The coronavirus disease 2019 (COVID-19) pandemic is one of the biggest healthcare challenges in history. As effective treatment options are lacking and several vaccines are already available, it has been estimated that a pandemic spread could be stopped if more than 67% of the population acquire immunity by either vaccination or infection, i.e. if herd immunity has developed (1). Allowing uncontrolled infection to achieve this immunity would cause increased morbidity and mortality and unjustifiable strain on the health system (2). This became apparent during the first and second waves of the pandemic, when even advanced health care systems were overrun by the surge of severely ill patients. Furthermore, even mild to moderate COVID-19 infections might lead to long-term health consequences (3) and individual financial losses (4). Therefore, large scale population-wide vaccination programs are the preferred approach to stop the pandemic and have been started worldwide. However, if there is a lack of a sufficient vaccine acceptance among the population herd immunity cannot be achieved. Vaccine hesitancy is a global barrier to this strategy and rated among the top ten threats worldwide (5, 6).

Consequently, even if governments can provide sufficient vaccine supply to vaccinate the total population (which already is a major challenge), herd immunity cannot be achieved if pronounced vaccine hesitancy is present (7). Health care workers (HCW) serve as important multiplier and advisor role in vaccination programs, as they are required to provide information and thus build confidence. Additionally, HCWs who are at risk of acquiring infections from their patients, are often regarded as the source of nosocomial infections with vaccine preventable diseases (8) - a circumstance that appears particularly threatening during the COVID-19 pandemic. In a representative national survey in Germany, the reported acceptance to receive a COVID-19 vaccination was only 66% in the general population (having risen from 50% in December 2020) and slightly lower with 64% in a small subset of HCW (9), **Table 1**. Recent, partially not yet peer-reviewed surveys of HCW have shown vaccine acceptance rates between 31% and 86% (10-24) (**Table 1**). Yet, these findings were the basis for negative media coverage in the German and international press shedding a picture of vaccine hesitancy among HCW. To design specific vaccination campaigns, addressing the concerns of potential vaccine candidates in the health care system, public health stakeholders need sufficient data reflecting the reasons for hesitancy. In the context of an ongoing pandemic caused by a new infectious disease, previous findings on vaccine hesitancy for classical vector-based vaccines such as annual influenza vaccines might not be completely applicable to COVID-19 vaccines. This is even more understandable since a new technology of mRNA-based vaccines are used for the first time. The individual decision to receive a COVID-19 vaccination is multifactorial, based among others on a individually perceived health risk, experiences with past vaccinations and sociodemographic factors (25).

**Table 1.** Overview on studies identifying vaccination acceptancy in medical professionals.

| Publication | Country | Participants total | Subgroups | Acceptance (% of participants Apr 20 2021) | Hurdles / Barriers | Peer-reviewed | Survey conducted |
| --- | --- | --- | --- | --- | --- | --- | --- |
| Holzmann-Littig et al. (present manuscript) | Germany | 4500 | HCW | 91.7% | development velocity, lack of trust in authorities, personal surrounding against vaccination, lack of information, generally not keeping vaccinations up to date | Preprint | Feb 21 |
| Wang et al. (10) | Hong Kong | 806 | Nurses | 40.3% | “suspicion on efficacy, effectiveness and safety”, “believing it unnecessary”, and “no time to take it” | yes | Feb / Mar 20 |
| Kwok et al. (11) | Hong Kong | 1205 | Nurses | 63.0% | 5 C | yes | Mar / Apr 20 |
| Dror et al. (12) | Israel | 211 | Nurses | 61.1% | No care of COVID-19 patients | yes | Mar / Apr 20 |
| Gagneux-Brunon (13) | France | 1554 | HCW | 76.9% | vaccine hesitancy | yes | Mar-Jul 20 |
| COSMO (9, 14) | Germany | 127 | n.a. | □ 64 % | Knowledge / information, trust | no | Dec 20 |
| Karagiannidis et al. (15) | Germany | 2305 | Physicians, Nurses | 64% |  | yes | Dec 20 |
| DIVI/DGIIN (16) | Germany | 3501 | Physicians, Nurses | 75% |  | no | Feb 21 |
| Grech et al. (17) | Malta | 123 | GP's and GP trainees | 61.8% | Knowledge, long-term adverse effects | yes | Sep 20 |
| American Nurses Foundation (18) | USA | 12939 | Nurses | 34.0% | Development to quick, not enough information, unclear clinical trials process | no | Oct 20 |
| Revive Health (19) | USA | 370 | Physicians, Nurses | 31.0% | Safety, efficacy, compromised development process | no | Oct / Nov 20 |
| Di Gennaro et al (20) | Italy | 1723 | HCW | 67% | Lack of trust in vaccine safety, little / conflicting information | yes | Oct/Nov 20 |
| Verger et al. (21) | France, Belgium, Quebec (Canada) | 2678 | HCW | 71.6% | Vaccine safety concerns | yes | Oct / Nov 20 |
| Papagiannis et al. (22) | Greece | 340 | HCW | 78.5% | fear of adverse effects, shortness of vaccine development time | yes | Dec 20 |
| Meyer et al. (23) | Pennsylvania, USA | 16158 | HCW | 55.0% | Unknown risks, insufficient data, adverse effects, wanting to wait to see how it goes with others | Preprint | Dec 20 |
| Schrading et al. (24) | USA | 1398 | HCW | 86.0% | Safety concerns | yes | Jan 21 |

Abbreviations: GP, general practitioner; DIVI, Deutsche Interdisziplinäre Vereinigung für Intensiv- und Notfallmedizin; DGIIN, Deutsche Gesellschaft für Internistische Intensivmedizin und Notfallmedizin; HCW, health care worker; n.a., not applicable.

The main goals of this survey in HCW were A) to provide current numbers of HCW COVID-19 vaccination acceptance in Germany, B) to identify vaccination barriers by assessing vaccine hesitancy on different scales. By identifying concerns and reasons for vaccination hesitancy, we aim to provide a data foundation for health stakeholders designing target group specific vaccination campaigns.

## Materials and Methods

In order to build tailored evidence syntheses on COVID-19 research for German health care professionals, politicians, stakeholders and laity, CEOsys (COVID-19 evidence ecosystem, www.covid-evidenz.de),(26) a research group within the network of German university hospitals (NUM, Netzwerk Universitätsmedizin) was launched in 2020. The network is funded by the German Federal Ministry of Education and Research (Bundesministerium für Bildung und Forschung, BMBF). This study was conducted as part of the evaluation of informational needs. The study adheres to the declaration of Helsinki. Approval by the local ethics committee of the medical faculty of Technical University of Munich (41/21 S), data protection officer, hospital board and staff counsel were obtained. Every participant gave informed consent by clicking a checkbox after the information on the study and data protection prior to the survey. This cross-sectional study is based on an online survey conducted from 02/02/2021 to 02/28/2021 in German language. At that point of time, the vaccines Comirnaty^®^ by BioNTech/Pfizer and Vaxzevria®, the vaccine by Astra Zeneca had been in use in Germany. The vaccination campaign had been started approximately one month before the start of this survey.

The link to this voluntary open online survey was sent to a total of 3924 e-mail addresses of nursing homes, medical practices, ambulance services, medical universities, hospitals, ambulatory care services and medical societies across Germany. In the invitation email, it was asked to forward the link within the respective institution or society. No further advertising was carried out by the authors. There were no incentives offered for taking part in the survey. The survey was carried out on the online platform SoSci Survey (27). The items were not randomized. To guarantee full anonymity, no cookies to identify unique users, no IP checks or logfile analyses were performed and users were not asked for registration. There was no review step for the respondents.

To formulate questions, the GESIS (Leibniz Institute for the Social Sciences) survey guidelines for question wording were considered (28). The recommendations of the Working Group on Vaccine Hesitancy Determinants Matrix (30) were applied. The matrix included contextual influences, individual and group influences as well as vaccine/vaccination-specific issues. In addition, the 5C-model by Betsch et al. was taken into account when constructing the questions (31). Furthermore, questions were adapted from the surveys by Larson et al. (32). Also, questions addressing specific previous results on vaccine hesitancy were included (33). The questions were then discussed in detail with a sociologist (BL). Survey pretest was performed by several members of the authors’ departments as well as of the CEOsys network. The questions were then revised according to their comments. Testers reported a completion time of approximately ten minutes, which was included in the study information.

We followed the Checklist for Reporting Results of Internet E-Surveys (CHERRIES) (29) (**Supplemental Table 1**).

The survey contained 55 items (12 screens). Demographic data, previous vaccination behavior, trust in vaccines / physicians / pharma industry and health politics as well as fear of adverse effects, assumptions on the consequences of the disease, knowledge about vaccines and information seeking behavior of the participants were queried. Four items on the participants’ knowledge on COVID-19 vaccinations were included as well. The full questionnaire can be found in **Supplemental Table 2**. In professional groups and work settings, free text comments (if “other” was chosen) were evaluated and participants were assigned to professional groups where possible. The categories “dental assisting personnel”, “dentist” and “science” were built from the free text comments. For statistical analysis, the professional group “other non-physician medical staff” was built from the categories “non-examined nurse”, “medical specialist”, “non-physician staff in the rescue service” and “trainee”. The group “physicians with specialist / personnel responsibility” was built from the groups “specialized physician”, “consultant physician” and “chief physician”. In the item “Sources of information,” a maximum of five answer options could be selected to identify the media that are essential to the participant. Questionnaires which had not been completed (i.e., which had been abandoned), questionnaires without informed consent and questionnaires with missing information on vaccination willingness / hesitancy were excluded from statistical analyses. In several questions, the answer option “no answer” was given to comply with data protection guidelines on the one hand and to avoid abandoning of the questionnaire on the other.

### Statistical analyses

Data are presented as absolute and relative frequencies. Number of missing values (“no answer”) for all items are presented in the Supplement. For group comparisons odds ratios with corresponding 95% confidence intervals are presented and/or chi-square tests were performed. All tests were performed two-sided and a significance level of 5% was used. Due to the exploratory nature of the study no adjustment for multiple testing was considered. As the survey was most likely answered in work settings, i.e. completion might have been interrupted (leading to longer answer times or early termination and restart of the survey with shorter response times), time stamps were not analyzed.

Unless otherwise specified, participants were summarized to be “willing / accepting” if they wanted to get or had already been vaccinated. Similarly, participants who indicated they were undecided, or unwilling were summarized as hesitant. German regions were defined according to the classification used by the Robert Koch Institute (34). Statistical analysis was performed using R, version 4.0.4 (R Foundation for statistical Computing, Vienna, Austria) and its libraries “*epitools”*, “*arsenal”*, “*sf”* and “*mapplots”*.

## Results

### Study population

A total of 5,448 participants started the online survey. 948 data sets had to be excluded due to incompleteness or missing consent. Finally, the dataset consisted of 4,500 completed surveys for analysis (**Figure 1**). The number of female participants was 2,610 (58.0%). The largest participant groups were physicians with specialist / personnel responsibilities (29.5%), medical students (29.2%), and certified nurses (10.4%). Main work settings were maximum care hospitals / university hospitals (42.6%), hospitals of other care levels (18.0%), and medical practices / medical care centers (20.2%). Of all participants, 35.2% were from southern, 28.1% from northern, 26.0% from western, and 8.4% from eastern Germany (**Table 2**).

**Figure 1.**
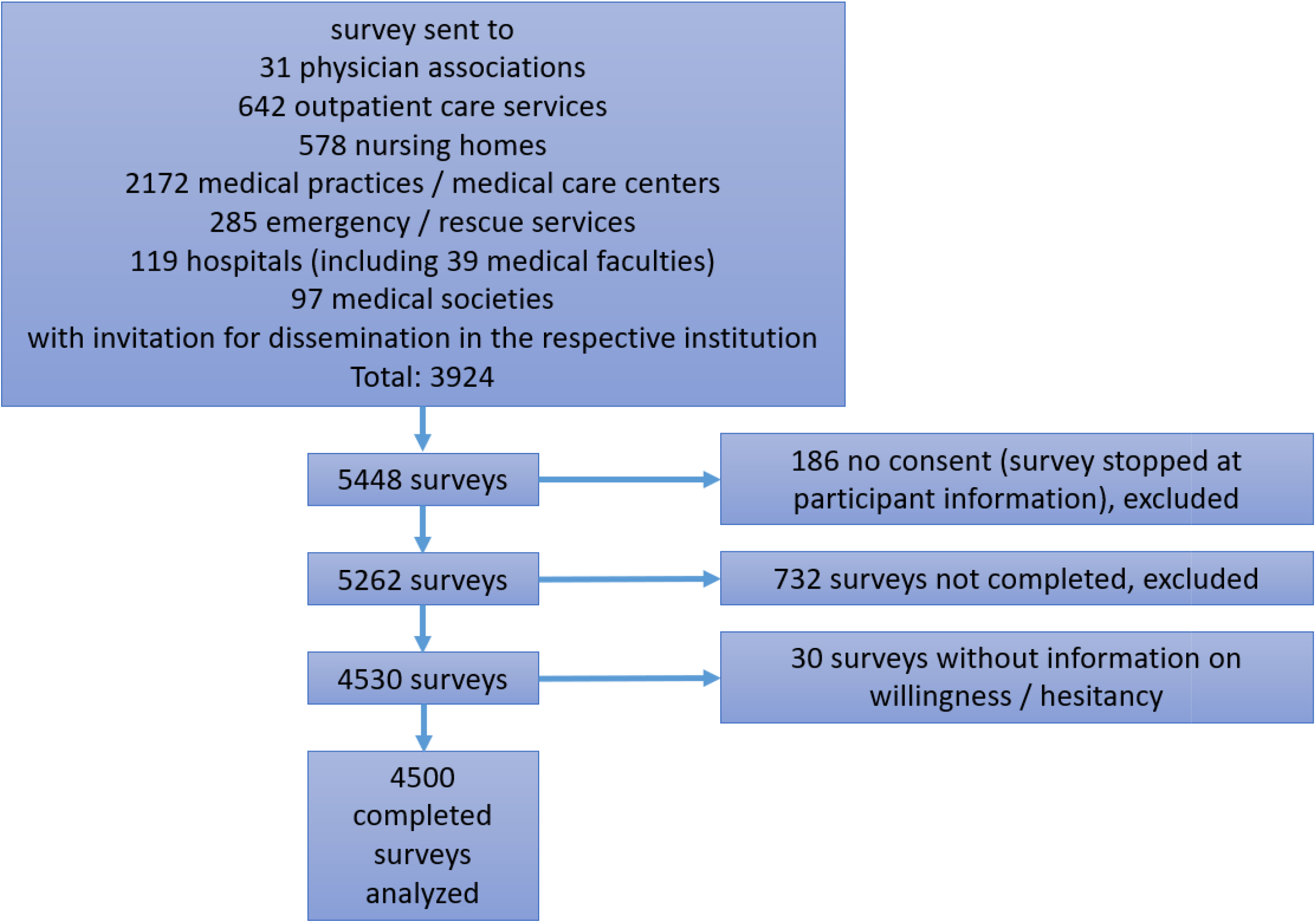
Flow-chart of participants.

**Table 2.**
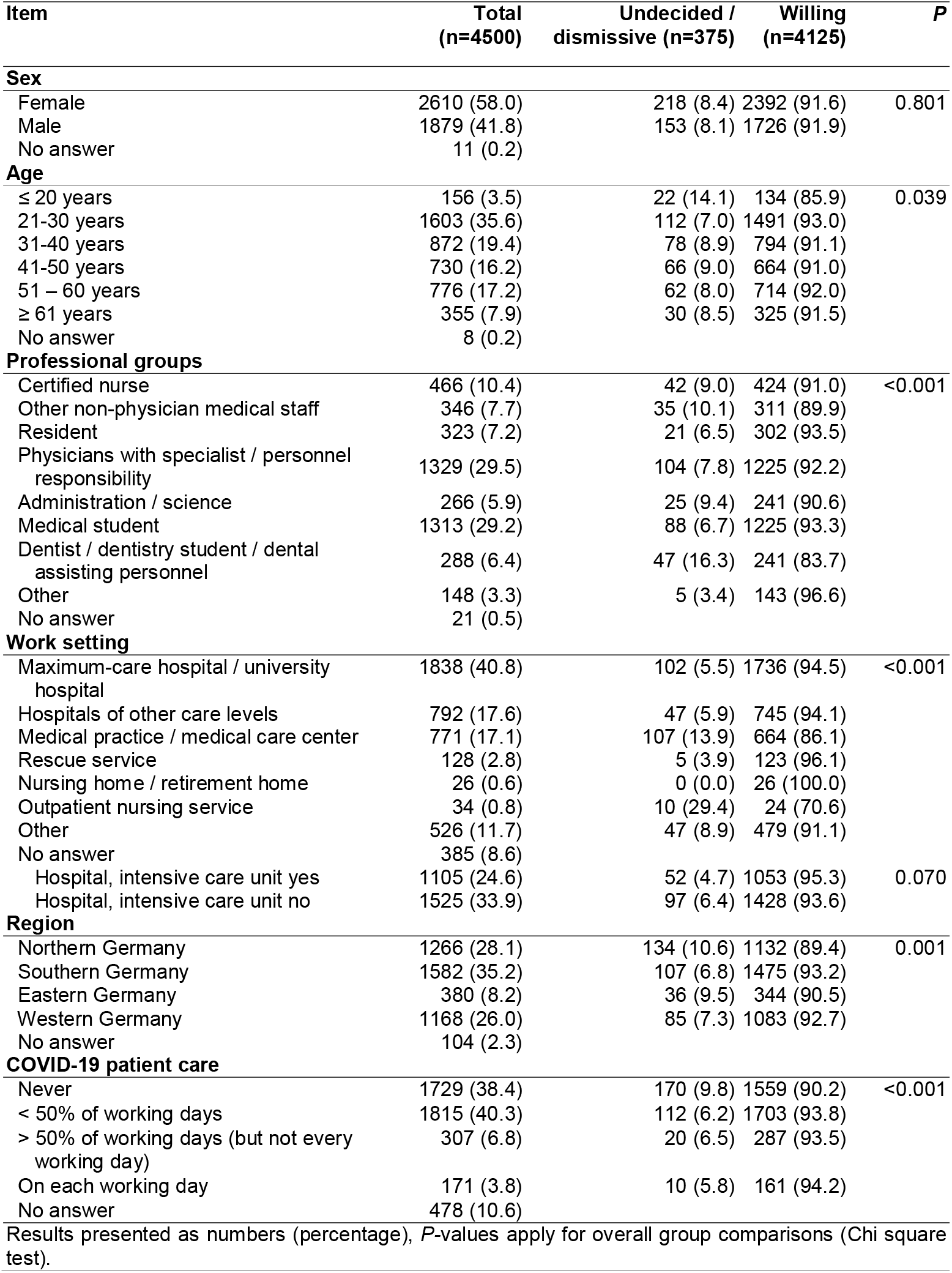
Overall study population, and differences between participants undecided/dismissive or willing to receive a COVID-19 vaccination.

The overall vaccination acceptance was 91.7% (4125 / 4500), 167 (3.7%) participants reported to be undecided, 208 (4.6%) reported that they do not want to get vaccinated against COVID-19. Of those accepting a COVID-19 vaccination, 2,024 (49.1%) had already received at least one COVID-19 vaccination dose. No difference in vaccination acceptance was observed between sexes (**Table 2**). Within German regions, the vaccination acceptance ranged from 89.4% (northern Germany) to 93.2% (southern Germany, **Figure 2**). Participants from outpatient nursing services (10 / 45, 22.2% unwilling or undecided) and medical practices / medical care centers (127 / 908, 14.0%) were less likely to accept a COVID-19 vaccination compared to participants from other work settings (**Table 2, Figure 3**). Vaccination acceptance varied significantly between age groups (p=0.039). The age group ≤20 years showed the lowest vaccination acceptance. (**Table 2, Figure 3**).

**Figure 2.**
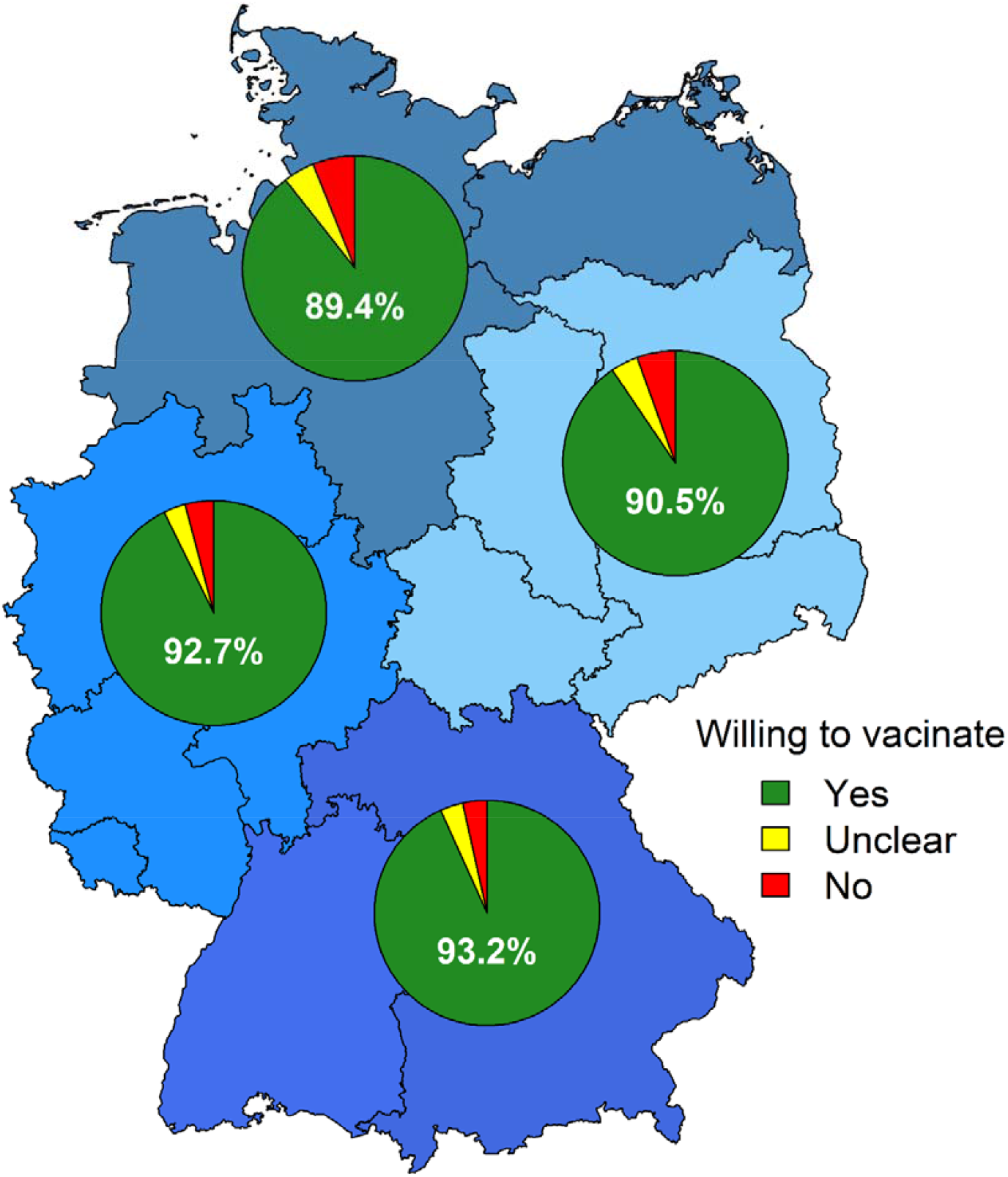
COVID-19 vaccination acceptance in German regions. Regions defined according to the region definitions in the COVIMO study.(34)

**Figure 3.**
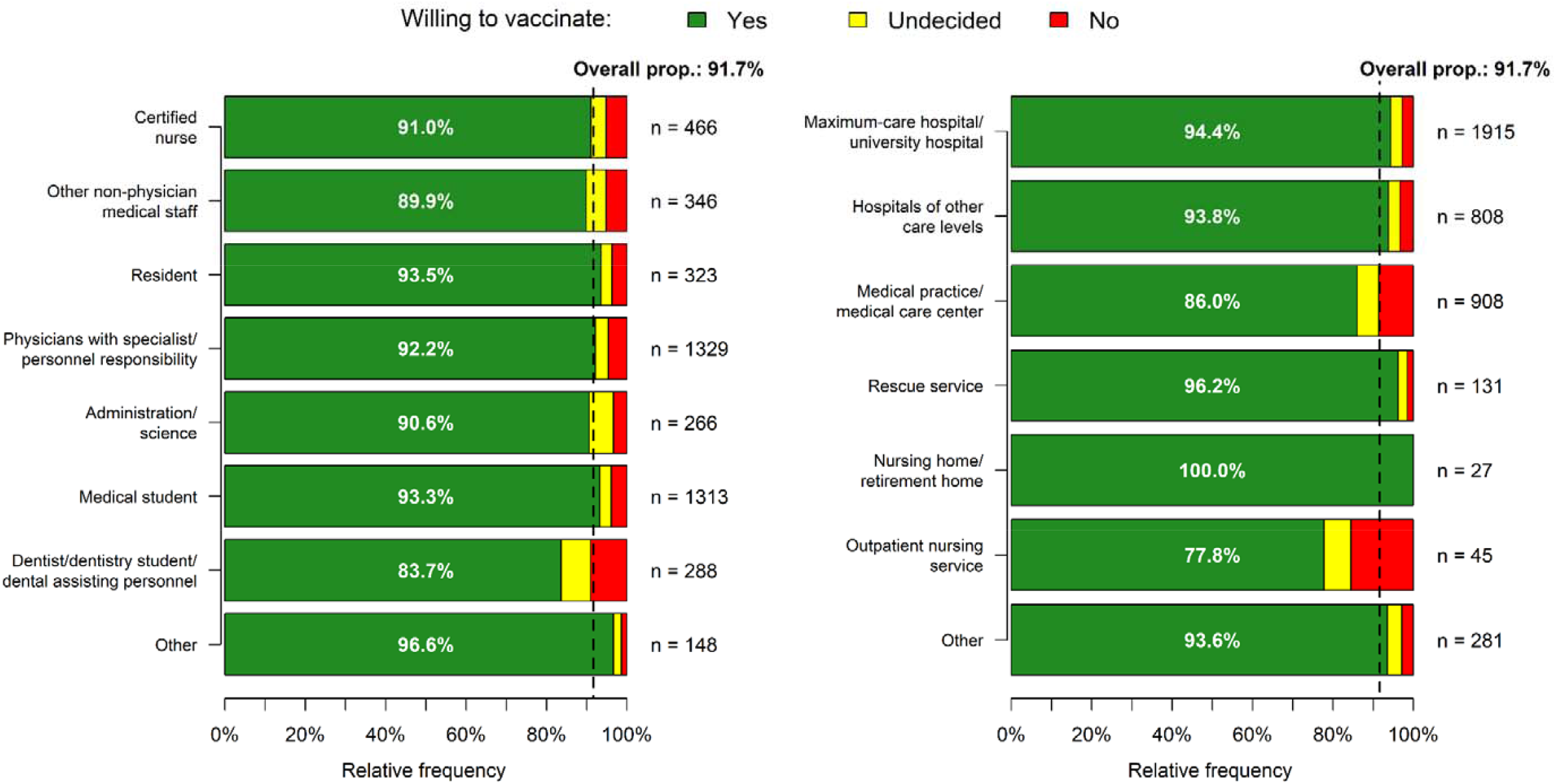
Vaccination acceptance in professional groups and work setting. The overall proportion of willing to vaccinate was 91.7%. Abbreviations: prop., proportion.

**Figure 4.**
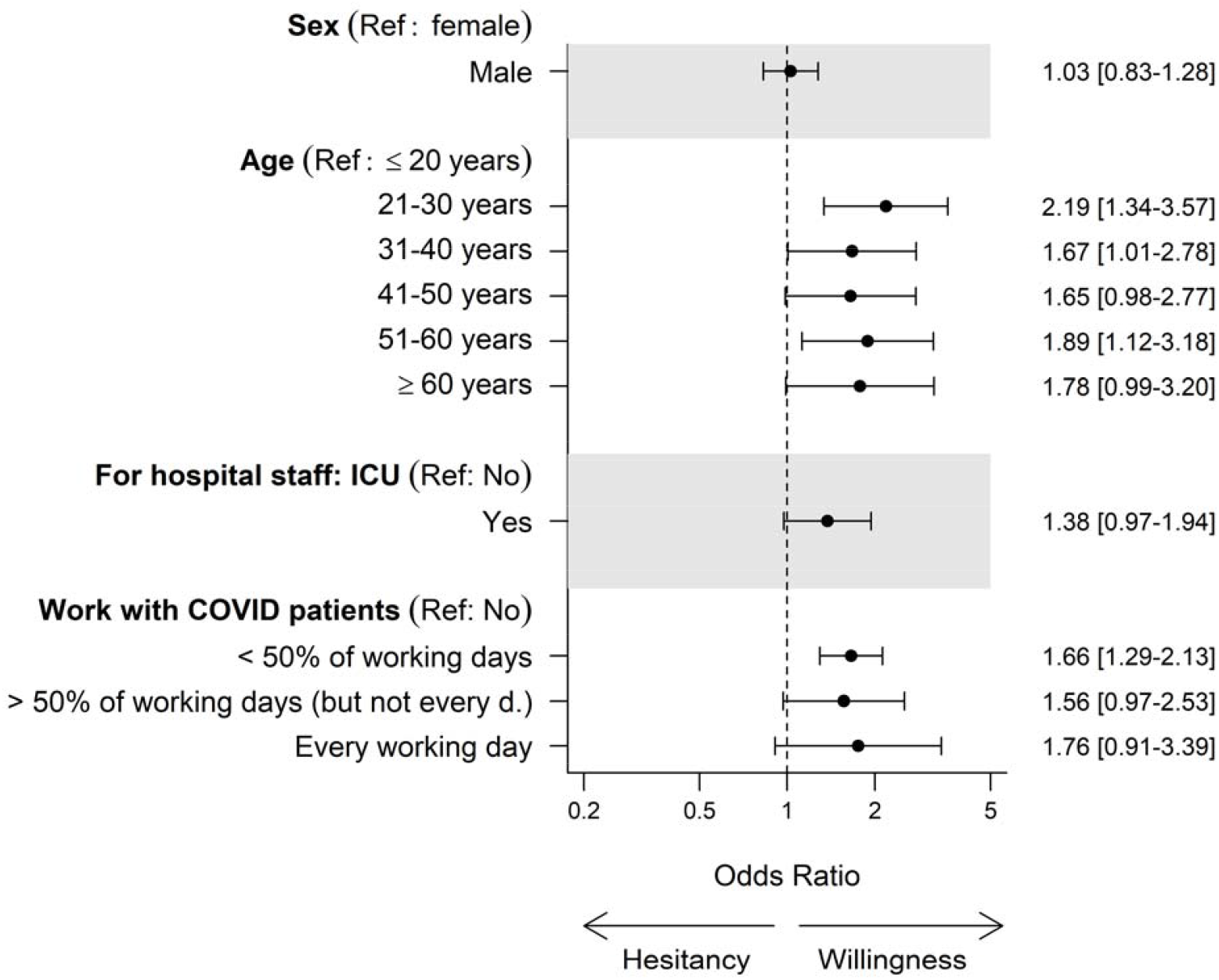
Basic demographics and vaccination acceptance. Abbreviations: Ref, reference; d., day; ICU, intensive care unit; COVID, coronavirus disease.

### Personal attitudes and vaccination experience

Overall, most participants rather or fully agreed that they trust both vaccines in general (4,199, 93.3%) and COVID-19 vaccines (3639, 80.9%). Most respondents rather or fully agreed that vaccines in general (4,330, 96.2%) and COVID-19 vaccines (3961, 88.0%) are effective. Additionally, most of the respondents fully or rather claimed to keep their vaccinations up to date (4150, 92.2%).

Lack of trust in regulatory authorities in general (115 / 198 [58.1%] of participants who totally disagreed or rather disagreed reported to be unwilling to get vaccinated or to be undecided versus 172 / 4,041 [4.3%] of those who rather agreed or totally agreed, p<0.001) and specifically in COVID-19 vaccine approval (181 / 322 [56.2%] vs. 85 / 3,763 [2.3%], p<0.001), as well as in German health politics (206 / 762 [27.0%] vs. 85 / 2,746 [3.1%], p<0.001), and physicians (51 / 164 [31.1%] vs. 224 / 3,740 [6.0%], p<0.001) were significantly associated with vaccine hesitancy (odds ratios for comparison with the reference group of those who neither agree nor disagree are illustrated in **Figure 5A**, absolute and relative frequencies for all categories are presented in **Supplemental Table 3**, odds ratios are presented in **Supplemental Table 4**). A history of vaccination adverse effects (30 / 136 [22.1%] vs. 325 / 4,323 [7.5%] of participants without, p<0.001), disagreement with generally keeping vaccines up to date (rather or totally disagree: 74 / 201 [36.8%], rather or totally agree: 269 / 4,150 [6.5%], p<0.001) and with regularly receiving vaccinations against influenza (275 / 1,463 [18.8%] vs. 77 / 2,713 [2.8%], p<0.001) were associated with COVID-19 vaccine hesitancy (**Figure 5B, Supplemental Table 3, Supplemental Table 4**). Yet, a strong association to COVID-19 vaccine hesitancy was found towards fear of long-term (rather or totally agree: 302 / 863 [35.0%], rather or totally disagree: 35 / 3,070 [1.1%], p<0.001) and short-term (159 / 596 [26.7%] vs. 160 / 3,377 [4.7%], p<0.001) adverse effects of COVID-19 vaccines, as well as vaccine adverse effects in general (133 / 469 [28.4%] vs. 176 / 3,705 [4.8%]). Disagreement on feeling well informed about vaccines in general (rather or totally disagree: 85 / 265 [32.1%], rather or totally agree: 219 / 3,824 [5.7%], p<0.001) and specifically on COVID-19 vaccines (174 / 398 [43.7%] vs. 135 / 3,649 [3.7%], p<0.001) was associated with vaccine hesitancy (**Figure 5C, Supplemental Table 3, Supplemental Table 4**). Further, increased vaccine hesitancy was found for participants who believe that financial profit is more important for the pharmaceutical industry than safety of their products in general (rather or totally agree: 197 / 663 [29.7%], rather or totally disagree: 78 / 2,713 [2.9%], p<0.001) and in the production of COVID-19 vaccines (214 / 609 [35.1%] vs. 76 / 2,854 [2.7%], p<0.001), or for participants who reported skepticism towards velocity of COVID-19 development (299 / 695 [43.0%] vs. 31 / 3,226 [1.0%], p<0.001), or the new mechanism of action (237 / 584 [40.6%] vs. 61 / 3,383 [1.8%], p<0.001) (**Figure 5C, 5D, Supplemental Table 3, Supplemental Table 4**).

**Figure 5.**
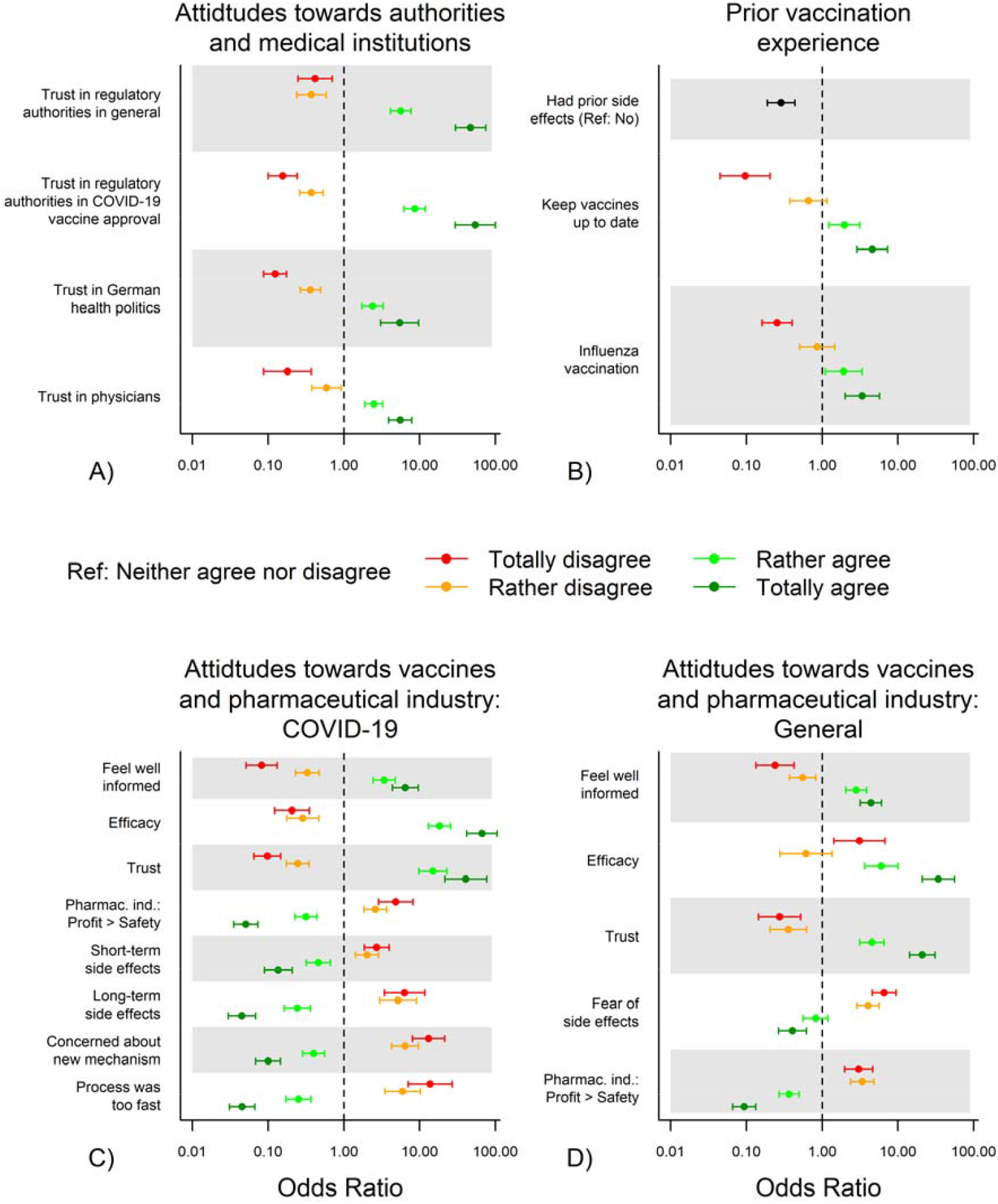
Attitudes and experiences with vaccinations. Attitudes towards authorities, and medical institutions (**A**), prior vaccination experience (**B**), towards COVID-19-specific vaccines and pharmaceutical industry (**C**), and generally towards vaccines and pharmaceutical industry (**D**). Abbreviations: Ref, reference; d., day; ICU, intensive care unit; incl., including; COVID-19, coronavirus disease 2019; Pharmac. ind., pharmaceutical industry

### COVID-19-specific attitudes and experiences in the personal surrounding

A strong association to COVID-19 vaccine hesitancy was observed, when participants stated that their personal surrounding had decided not to get vaccinated (183 / 251, 72.9%) compared to 59 / 3,682 (1.6%) for those who reported that the majority of their family/friends have or would like to have COVID-19 vaccination (p<0.001), and when their general practitioner had advised against a COVID-19 vaccination (30 / 41 [73.2%] vs. 18 / 518 [3.5%] with advise for vaccination, p<0.001). Also, a strong association between colleagues’ decisions against a COVID-19 vaccination and hesitancy was observed (not get vaccinated: 106 / 173 [61.3%], get vaccinated: 116 / 3,602 [3.2%], p<0.001). If participants did not know whether persons in their acquaintances had already been vaccinated against COVID-19 (11 / 56, 19.6%) or if none of these had been vaccinated (147 / 948, 15.5%) relevant vaccination hesitancy was present (217 / 3,495 [6.2%] for participants who reported that people in the personal environment already received at least one vaccination dose), (both p<0.001). Participants’ fear of an infection in the professional environment (rather or totally agree: 78 / 2,307 [3.4%], rather or totally disagree: 248 / 1,451 [17.1%], p<0.001) was stronger associated to vaccine acceptance/hesitancy than their fear of an infection in the private environment (32 / 1,387 [2.3%] vs. 297 / 2,225 [13.3%]) (**Figure 6, Supplemental Table 3)**.

**Figure 6.**
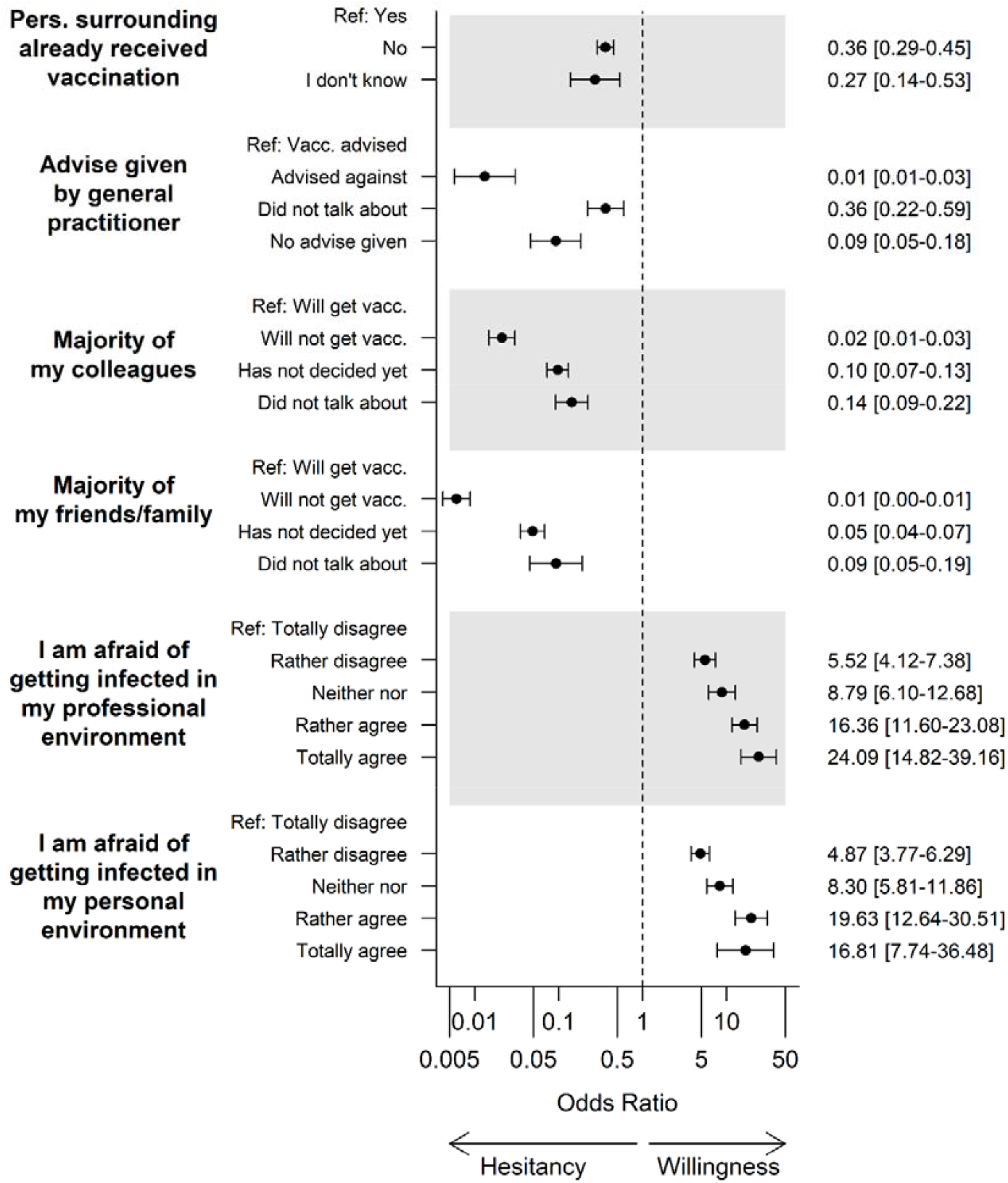
Personal environment and attitude towards COVID-19 vaccination. Abbreviations: Vacc., vaccination; GP, general practitioner; Pers, persons; Ref, Reference.

In participants expecting a severe COVID-19 disease course in individuals in their personal environment, the number of hesitant or undecided participants was 141 / 3,197 (4.4%) vs 117 / 655 (17.9%) of those not expecting this (p<0.001), **Figure 7**. Participants who felt that they were at risk of a severe disease course were significantly less often hesitant than participants not fearing a risk of a severe course of the disease for themselves (rather or totally agree: 30 / 941 [3.2%], rather or totally disagree: 292 / 2,658 [11.0%], p<0.001). Only small differences in vaccination acceptance/hesitancy were observed with regard to the history of COVID-19 infections, hospitalizations, intensive care unit admissions, and deaths in the participants’ acquaintances. Number of correct answers in the knowledge test was significantly associated with frequency of vaccine hesitation (zero correct answers: 31 / 94, 33.0%, all four questions answered correctly: 64 / 2,224, 2.9%, p<0.001) (**Figure 7, Supplemental Table 3**).

**Figure 7.**
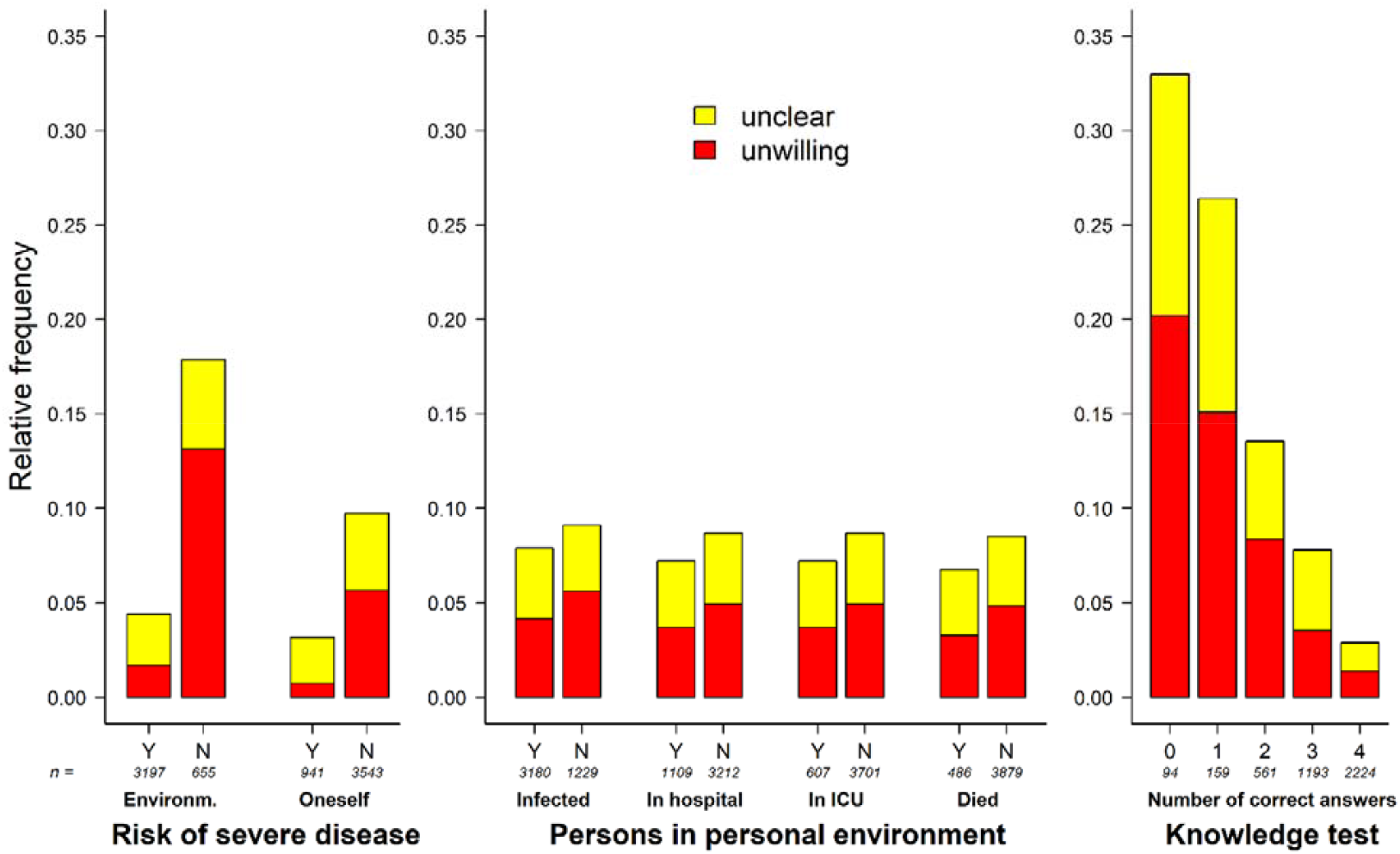
Perceived risk of a severe COVID-19 disease, history of infections in the personal environment and knowledge on COVID-19 vaccines. Abbreviations: ICU, intensive care unit; environm., environment; Y, Yes; N, No. Risk of severe disease for oneself (totally agree to totally disagree) was dichotomized for visualization. Category “I do not know” was not visualized. All data are given in Supplemental Table 3.

### Main Sources of Information on COVID-19

In the group of vaccine hesitant participants, lower proportions reported to obtain COVID-19-vaccination related information from online newspapers (42.4% vs 58.0%, p<0.001), TV / radio (46.9% vs. 60.8%, p<0.001), or websites / media of federal agencies (63.7% vs 76.0%, p<0.001). Furthermore, in this group, it was more frequently reported that COVID-19-vaccination information was obtained via messenger services (13.6% vs 4.4%, p<0.001) or online video platforms (23.7% vs 12.1%, p<0.001) (**Figure 8**).

**Figure 8.**
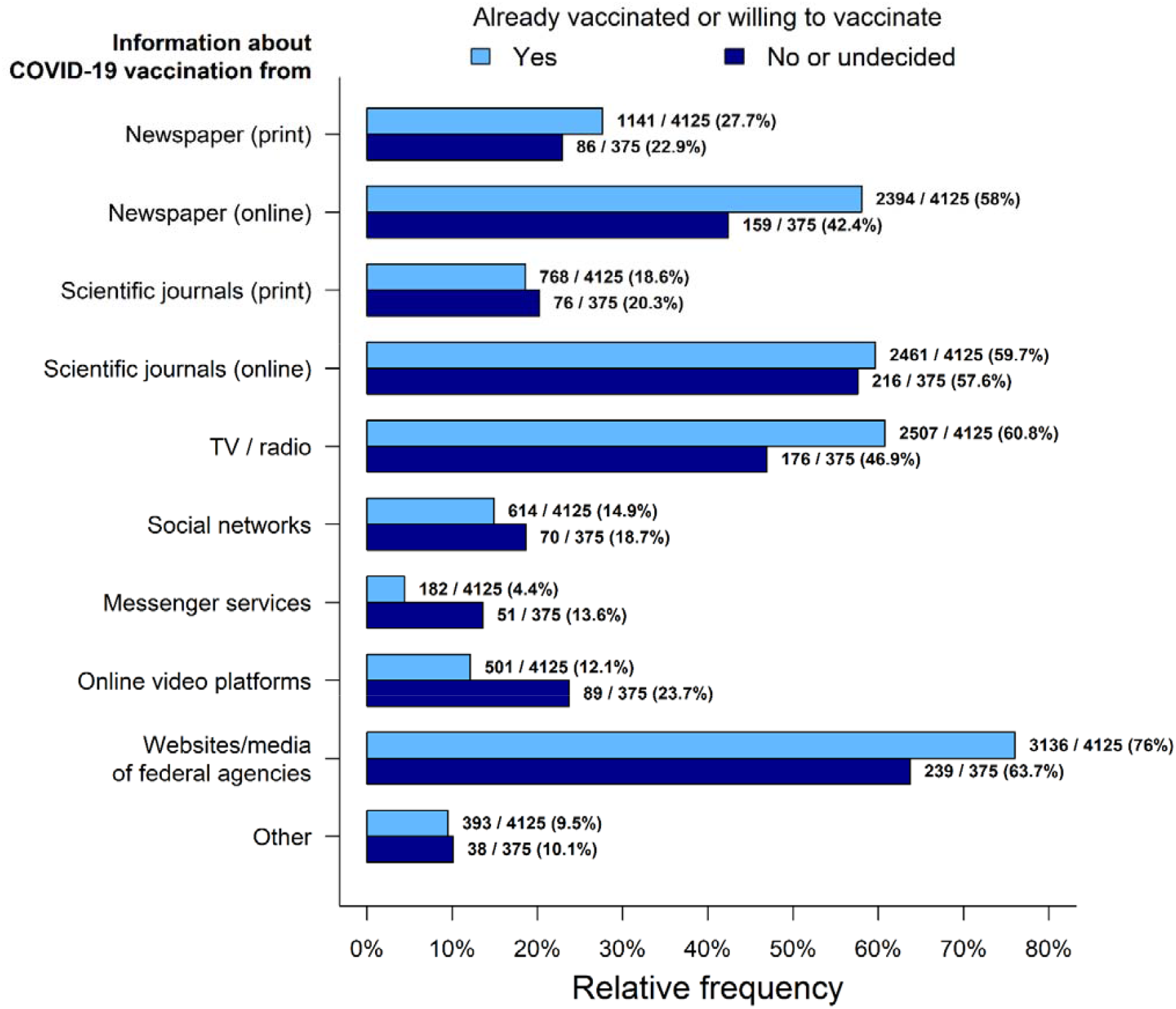
Source of information and media usage

## Discussion

We examined COVID-19 vaccine hesitancy in German health care workers (HCW) in one of the largest and most comprehensive surveys. Overall, data of 4,500 German HCW who participated in the survey during the second pandemic wave in Germany could be considered for this analysis. Of these, about 92% were either willing or already had received the vaccination. This high percentage of acceptance contrasts with recently published studies in Germany and worldwide (see **Table 1**) and shows that vaccine hesitancy may change rapidly. We could furthermore show that vaccine hesitancy in HCW is multifactorial. The identified factors associated with COVID-19 vaccination hesitancy might help to shape specific vaccination campaigns.

In this pandemic, vaccine hesitancy among HCW is an urgent issue (7) that needs to be addressed rapidly. Comparison to previously published studies as shown in **Table 1** needs to be done cautiously as acceptance was defined differently, measured on different scales, using different questionnaires with different numbers of items asked in different countries and time periods. One strength of this study is that the survey was conducted after the Comirnaty^®^ vaccine by BioNTech/Pfizer had been approved and was available for German HCW in hospitals. HCW are ranked priority in a situation with limited availability of vaccines to maintain critical health care functions which are especially needed in a pandemic situation. As this group of people has the highest exposure and risk of disease transmission, they had a realistic chance of getting vaccinated very soon in comparison to the hypothetical question asked in other surveys where no vaccination had been approved by health care authorities at the time of the survey. Online surveys are a powerful tool to achieve a broad response in short periods of time and therefore especially of use in a pandemic situation (35). This is opposed to conventional methods with pen-and-paper and careful selection of a representative sample which would simply take too much time in extremely fast changing circumstances (35). In addition, an online survey already seemed more than advisable given the distance requirements and the necessary reduction of interpersonal contacts.

### Vaccine hesitancy in health care workers

General vaccine hesitancy in HCW is common (8, 36) and recently published surveys observed substantial skepticism towards receiving a COVID-19 vaccination (summarized in **Table 1**). Previously, COVID-19 acceptance rates had been observed to range between 31% and 86%. The only comparable German survey was conducted mainly in intensive care settings. In that study, the acceptance of 50% in nurses and 73% in physicians was rather low compared to our data (15). However, the earlier assessment in December 2020 with an absence of approved and available vaccines could explain these differences. These low numbers would be insufficient to reach herd immunity in the medical sector (1). With 92% acceptance, we found a broad consent to receive a COVID-19 vaccination throughout all groups of HCW. As our survey has taken place in February 2021 and, therefore, much later than the previously published studies it is likely that the acceptance has changed during the pandemic, with co-health-care-workers already being vaccinated. As vaccination has started in January 2021 in most German hospitals, 44% of the participants in our study had already been vaccinated.

Also, compared with an estimated worldwide acceptance of around 70%, German HCW showed higher acceptance rates (25). This highlights again the high acceptance rate of German HCW but also points to substantial regional and cross-country differences that might not be generalizable. HCW often function as ambassadors for vaccine acceptance in the general population (36, 37). Therefore, our observations might lead to the conclusion that German HCW might have already acted as role models for vaccine acceptancy during the COVID-19 pandemic for their own peer group.

In Germany, influenza vaccination is recommended to all HCW (38). In our survey only 60% followed that recommendation, yet this rate was still higher than in some other published surveys in HCW (38, 39). This suggests a large-scale general vaccine acceptance in participating HCW which is in line with the observed general trust in vaccines, their efficacy, and the high agreement with the statement to keep vaccinations up to date. The perceived severe threat by COVID-19 might explain the higher acceptance to vaccinate against COVID-19 as compared to the seasonal influenza.

### Comparison of vaccine hesitancy in the German health care workers and the general population

The acceptance to receive a vaccination in the general population in Germany is measured weekly by the COSMO study (40). In that study, acceptance rates are relatively stable at 68% and therefore lower than in our survey (14). The ongoing debate about Astra Zeneca’s vaccine Vaxzevria®, can only serve to a limited extent as an explanation for this discrepancy, since this debate had just begun during our survey. However, it seems conceivable that the threat from COVID-19 appears less abstract for HCW than for the general population, which could lead to a higher willingness to vaccinate. Yet, preliminary results of the COVIMO study did not find major differences between German HCW and the general population regarding acceptance to receive a COVID-19 vaccine (preliminary results published online only. (34). However, the HCWs surveyed there were only a small subgroup.

### Factors influencing COVID-19 vaccine hesitancy

As media reports on vaccine hesitancy in German HCW claimed low acceptance rates, it became particularly important to assess the actual vaccination acceptance of HCW and reasons for hesitancy. In our study, 3.7% of all participants were undecided and 4.6% refused a COVID-19 vaccination. The subgroup of participants working in outpatient nursing services was stronger associated with a COVID-19 vaccination hesitancy with the caveat of a low number of participants in this subgroup. This might be due to the fact that most vaccine campaigns took place in hospitals and by health care providers, leading to the assumption that these workers were better informed. Working with COVID-19 patients was not associated with acceptance to receive a COVID-19 vaccine, except in HCW with less than 50% occupation with COVID-19 patients. This might be explained by the consideration of a perceived higher transmission risk in the family setting than in the professional setting where personal protective equipment and strict testing of patients and staff is used routinely, especially in COVID-19 treatment units, where HCWs treat COVID-19 patients on a daily basis. Also, it could be speculated that HCW in German hospitals were sufficiently supplied with professional protective equipment and therefore the threat of becoming infected with COVID-19 was judged lower (41).

The number of correct answers in the four COVID-19 vaccine knowledge questions was significantly associated with vaccination acceptance providing a direct starting point for informational campaigns. To test whether improving knowledge about COVID-19 vaccines is a modifiable risk factor leading to higher vaccination acceptance is recommended for further studies.

It is not surprising that prior adverse effects to vaccines went along with lower vaccine acceptance. The same could be seen for the factor of “up to date” vaccination and the acceptance to receive an influenza shot. This is in good accordance with previous studies (42). A general and COVID-19 vaccine-specific mistrust in the pharmaceutical industry was also associated with COVID-19 vaccine hesitancy.

### Possible implications for COVID-19 vaccination campaigns

Overall, the acceptance in HCW was remarkably high throughout Germany with a tendency of lower acceptance in northern and eastern parts of Germany. However, this small difference might not justify applying different vaccine campaigns in different German regions. Due to the high acceptance rate in HCW, they should therefore be involved as ambassadors for the COVID-19 vaccination campaign. We suppose that HCW already serve as a role model. Inclusion of HCW in vaccination campaigns might contribute to improved trust in the vaccination. It seems that low numbers of hesitant HCW have high distrust in politicians and health authorities. Having identified trust in vaccines and authorities as a potential modifiable risk factors in HCW, this might not be changeable in short-term campaigns but important in long-term projects. As hesitant HCW are using more streaming video platforms and messenger services it might be reasonable to use these media in designing a vaccination campaign. We assume that the produced content should be of high-level quality in regard to content and storytelling. This implies the need of multi-professional teams including medical education media professionals. As knowledge about the COVID-19 vaccine is a strong factor in favor of the vaccination, “infotainment” formats, e.g., easy to understand explanatory videos might be useful. Importantly, to evaluate the effect of COVID-19 vaccination campaigns a scientific supervision appears to be advisable.

Limitations of the study also need to be considered. As this in an open online survey it is impossible to estimate how representative the sample is. Due to the broad distribution of the survey without addressing specific individuals (due to German data protection laws) we cannot estimate how many HCW received the invitation to participate in the survey. Therefore, the study could be biased towards participants with a positive attitude towards a COVID-19 vaccination. Especially, mistrust in the state and organizations could further hinder participation. Furthermore, we have solely evaluated factors associated with vaccine hesitancy in HCW. Therefore, implications may primarily be drawn for this target group and not be generalizable to the general public. Finally, the small numbers in some subgroups allow only limited conclusions for these subgroups.

## Conclusions

In conclusion we saw a high overall acceptance rate amongst HCW to receive a COVID-19 vaccine. Furthermore, several factors associated with vaccination hesitancy in HCW were identified. Absence of knowledge about the vaccines and their mechanism of action as well as mistrust in authorities have been revealed as most important and potentially modifiable factors of vaccination hesitancy. HCW, due to their knowledge and overall high acceptance towards COVID-19 vaccination may prove as promising ambassadors for supporting the COVID-19 vaccination campaign. These factors could be addressed by public health stakeholders to assure a prompt achievement of herd immunity.

## Data Availability

Aggregated data is provided in the supplement. Due to German data protection law and local data protection guidelines, the full dataset may not be opend to the public. However, further data may be requested from the authors.

## Declarations

### Author Contributions

CHL, CSC, MP and BL designed the survey. CS, FF and JC reviewed the study design. CHL and CSC drafted the initial version of the manuscript. MCB revised the initial version and gave important intellectual content. CA, TF and JJM revised the manuscript and gave important advice on the interpretation of the results. BH performed the statistical analysis. All authors contributed important intellectual content and approved the final version of the manuscript.

### Ethics approval and consent to participate

The study adheres to the declaration of Helsinki. Approval of the local ethics committee of the Technical University of Munich School of Medicine (41/21 S), data protection officer, hospital board and staff counsel were obtained. Every participant gave informed consent by clicking a checkbox after the information on the study and data protection prior to the survey.

### Data Availability Statement

The datasets for this manuscript are not publicly available because written informed consent excluded data sharing as advised by the local data protection officer in accordance with the German data protection law.

### Code availability

Not applicable

### Consent for publication

Not applicable

## Supplemental Material

**Supplemental Table 1:** Appliance of the CHERRIES checklist, table adapted from the Cherries checklist (29)

| Item Category | Reported |
| --- | --- |
| **Design** |  |
| Describe survey design | √ |
| **IRB (Institutional Review Board) approval and informed consent process** |  |
| IRB approval | √ |
| Informed consent | √ |
| Data protection | √ |
| **Development and pre-testing** |  |
| Development and testing | √ |
| **Recruitment process and description of the sample having access to the questionnaire** |  |
| Open survey versus closed survey | √ |
| Contact mode | √ |
| Advertising the survey | √ |
| **Survey administration** |  |
| Web/E-mail | √ |
| Context | √ |
| Mandatory/voluntary | √ |
| Incentives | √ |
| Time/Date | √ |
| Randomization of items or questionnaires | √ |
| Adaptive questioning | √ |
| Number of Items | √ |
| Number of screens (pages) | √ |
| Completeness check | √ |
| Review step | √ |
| **Response rates** |  |
| Unique site visitor | n/a |
| View rate (Ratio of unique survey visitors/unique site visitors) | n/a |
| Participation rate (Ratio of unique visitors who agreed to participate/unique first survey | n/a |
| Completion rate (Ratio of users who finished the survey/users who agreed to participate) | √ |
| **Preventing multiple entries from the same individual** |  |
| Cookies used | √ |
| IP check | √ |
| Log file analysis | ✓ |
| Registration | ✓ |
| **Analysis** |  |
| Handling of incomplete questionnaires | ✓ |
| Questionnaires submitted with an atypical timestamp | ✓ |
| Statistical correction | ✓ |

**Supplemental Table 2.**
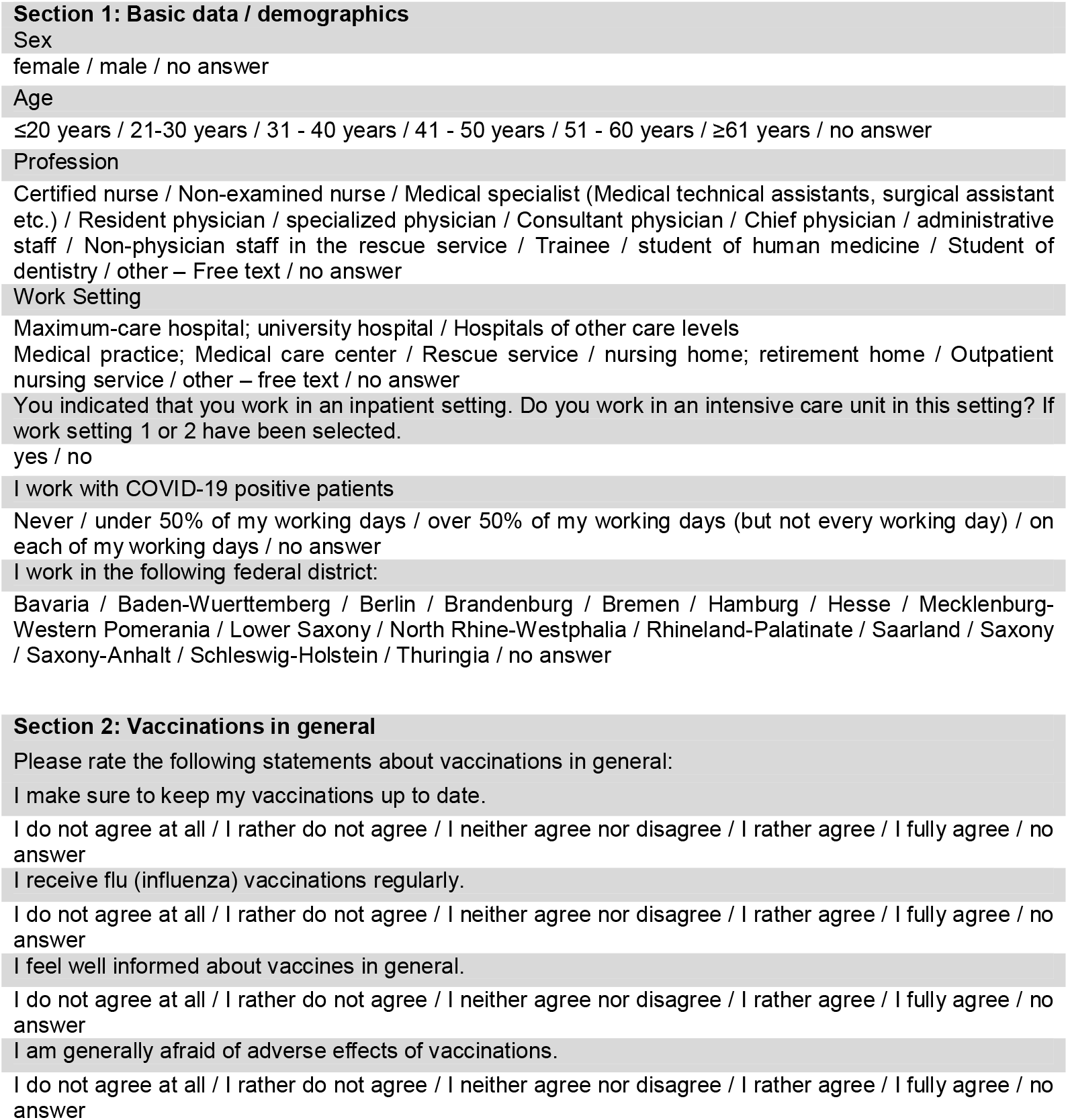
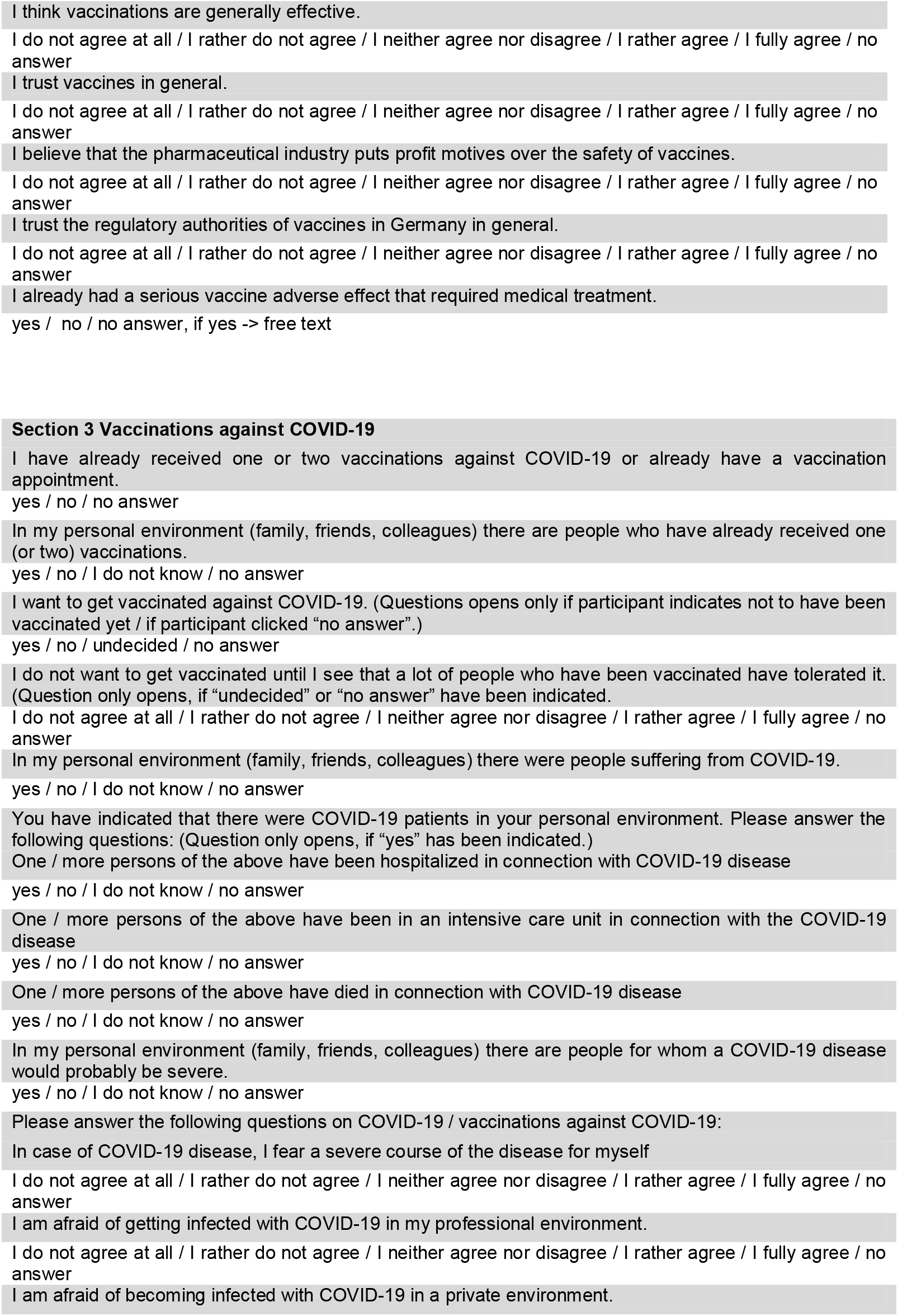
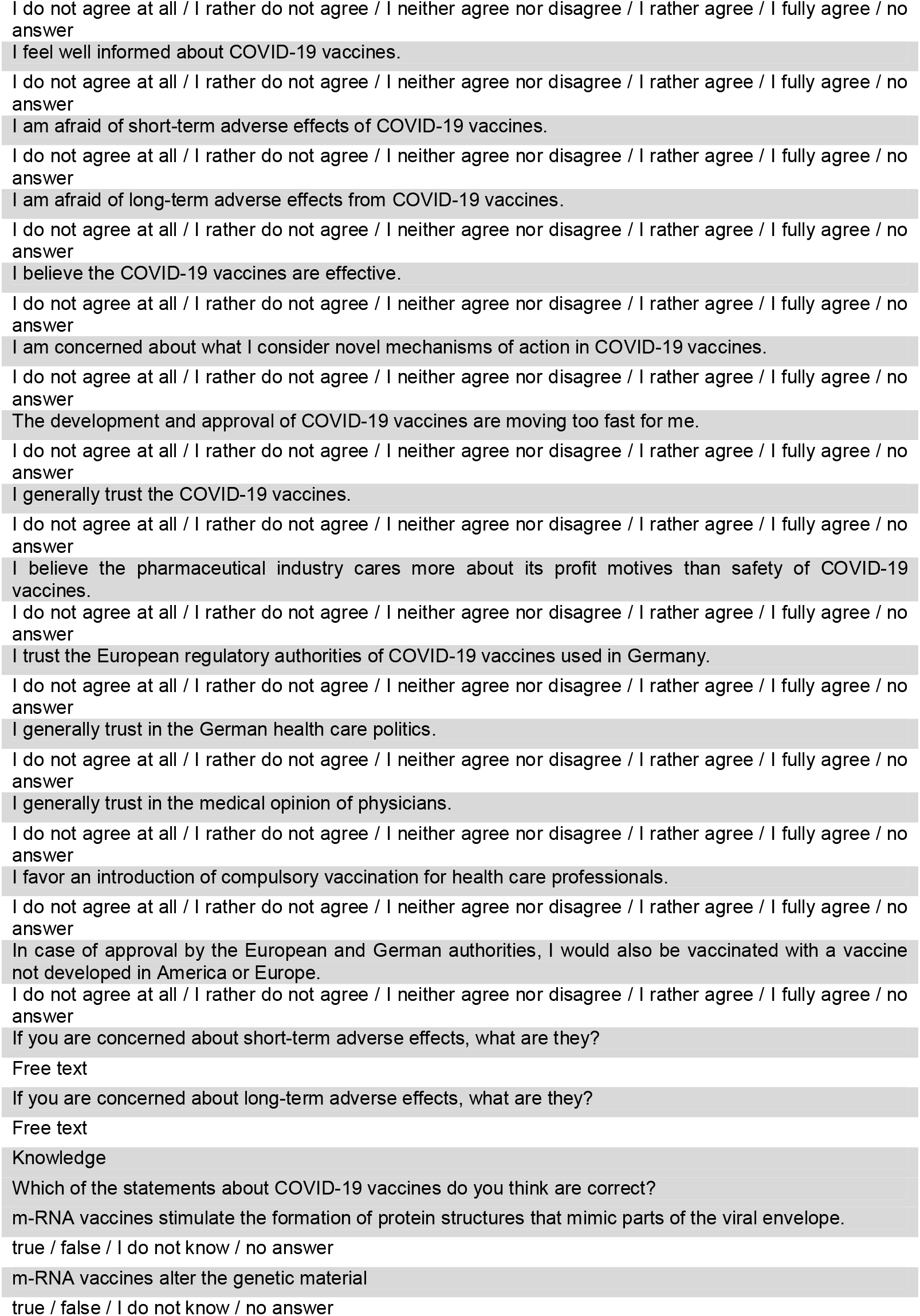
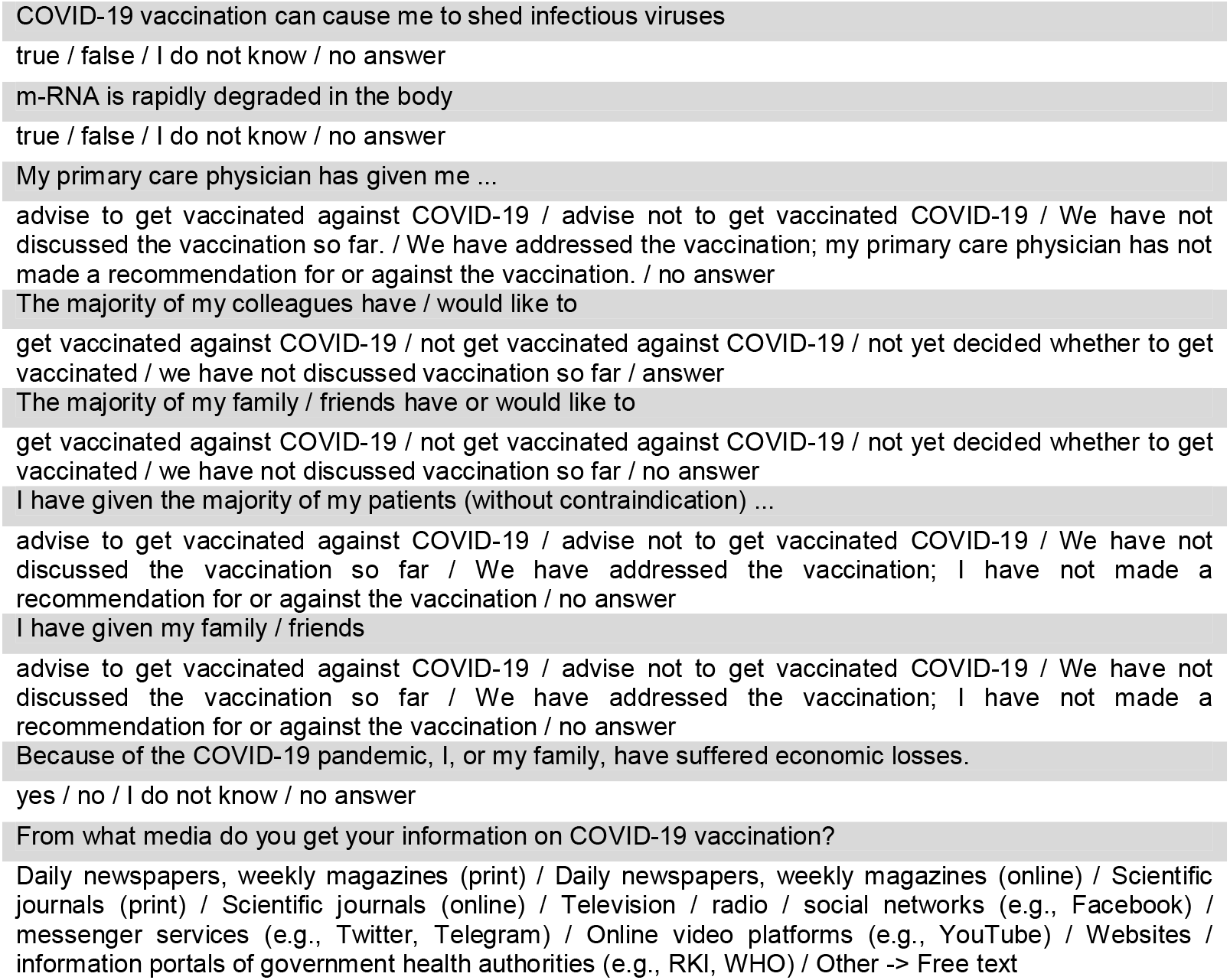
Original questions of the survey, translated from German.

**Supplemental Table 3:**
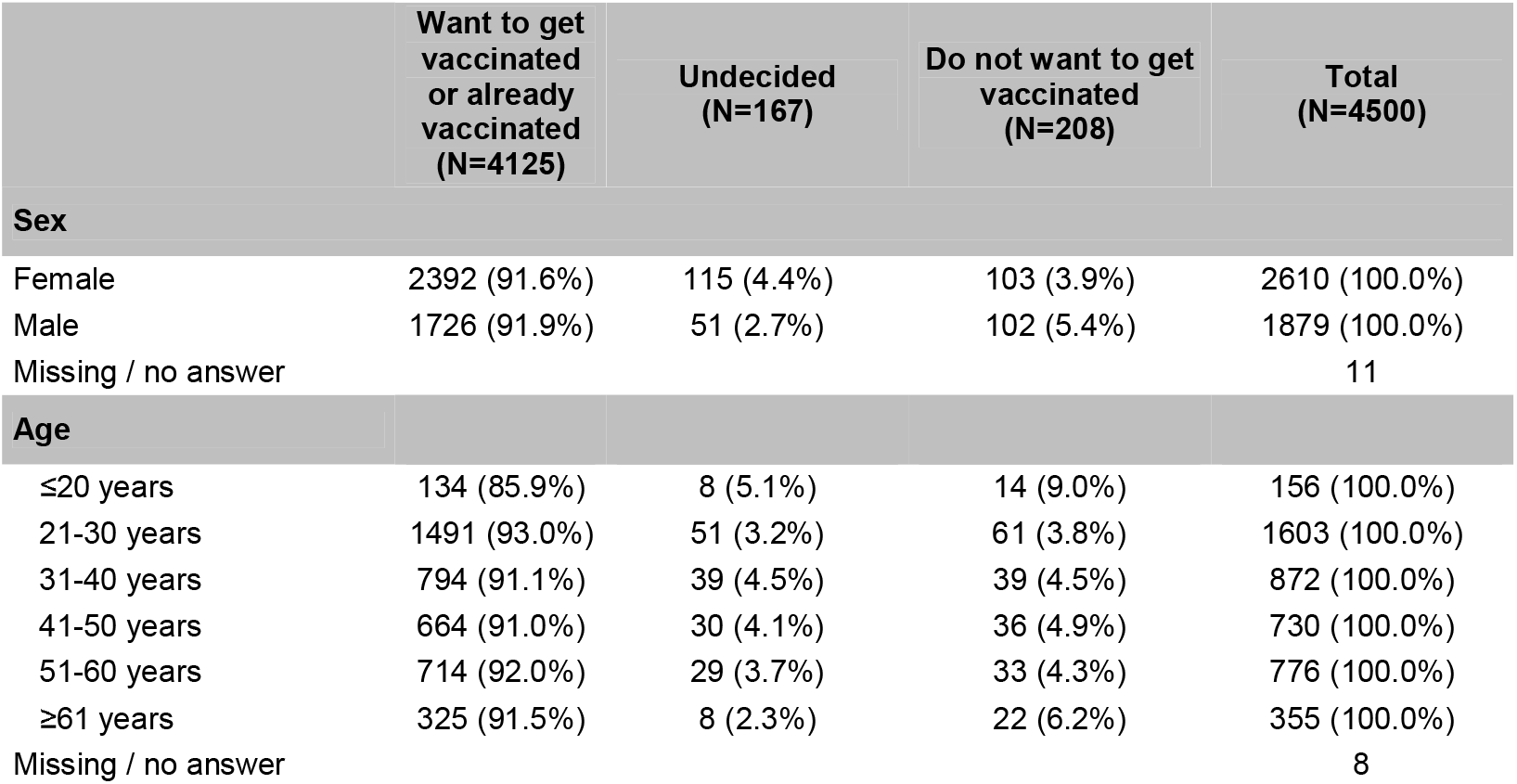
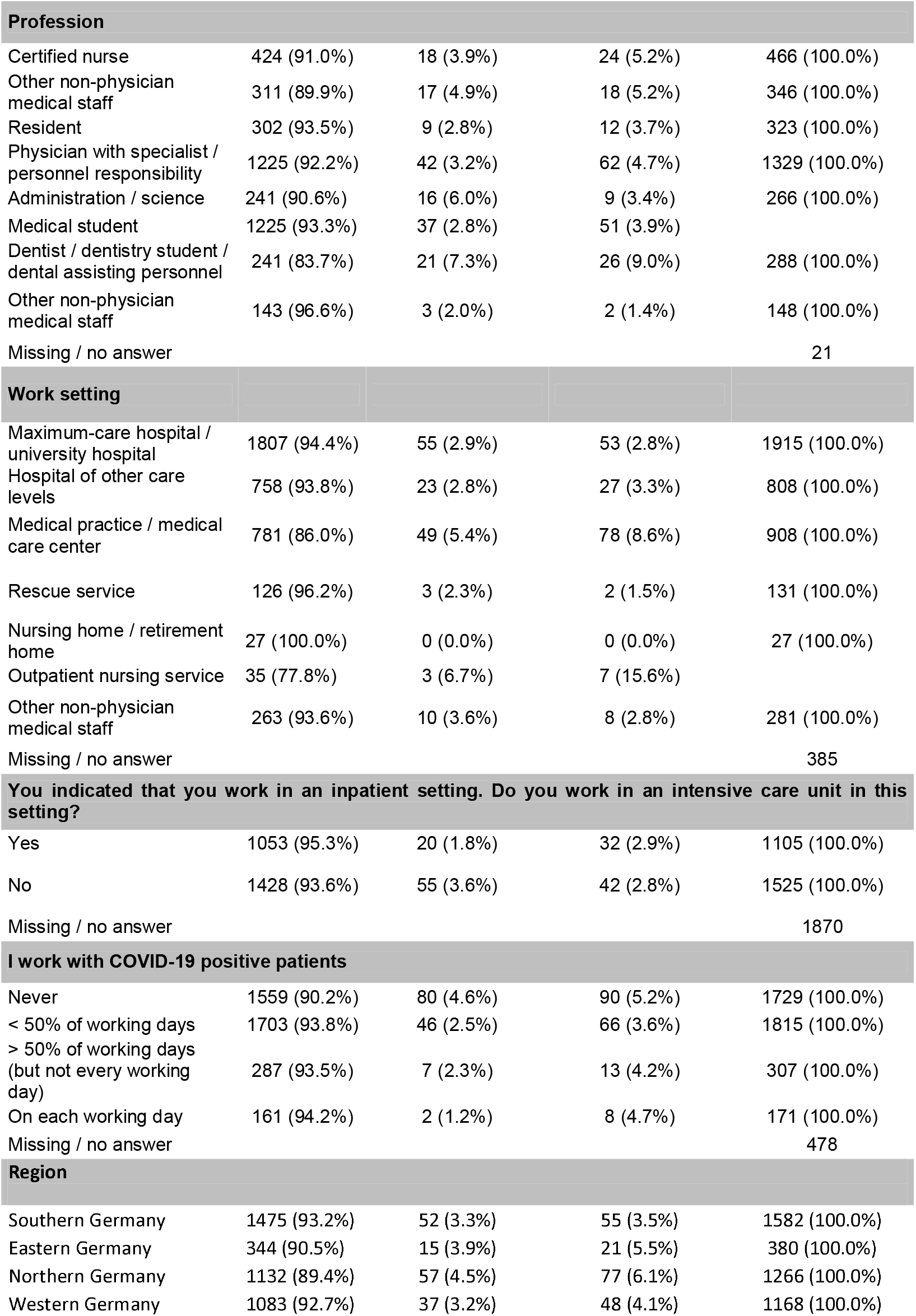
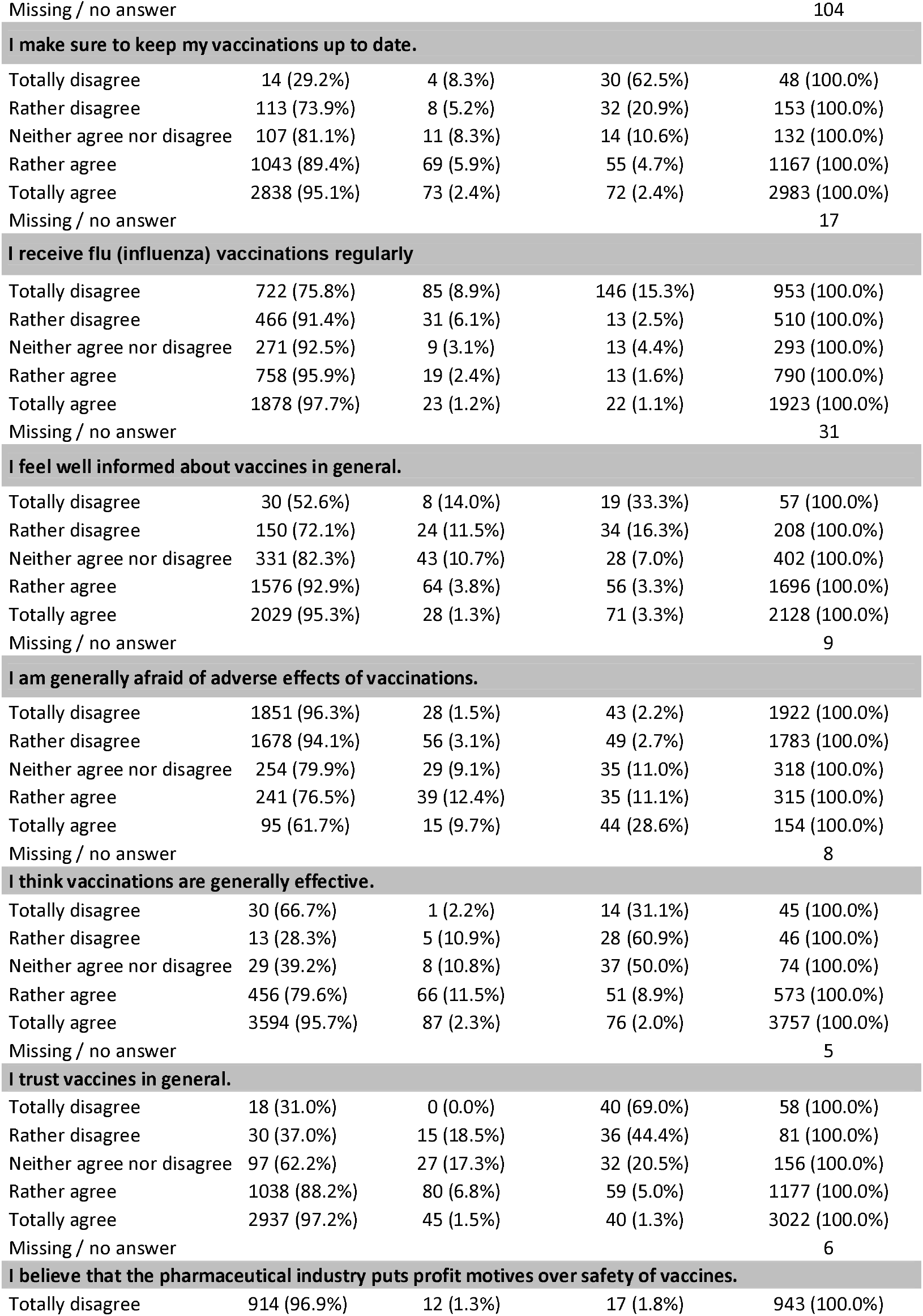
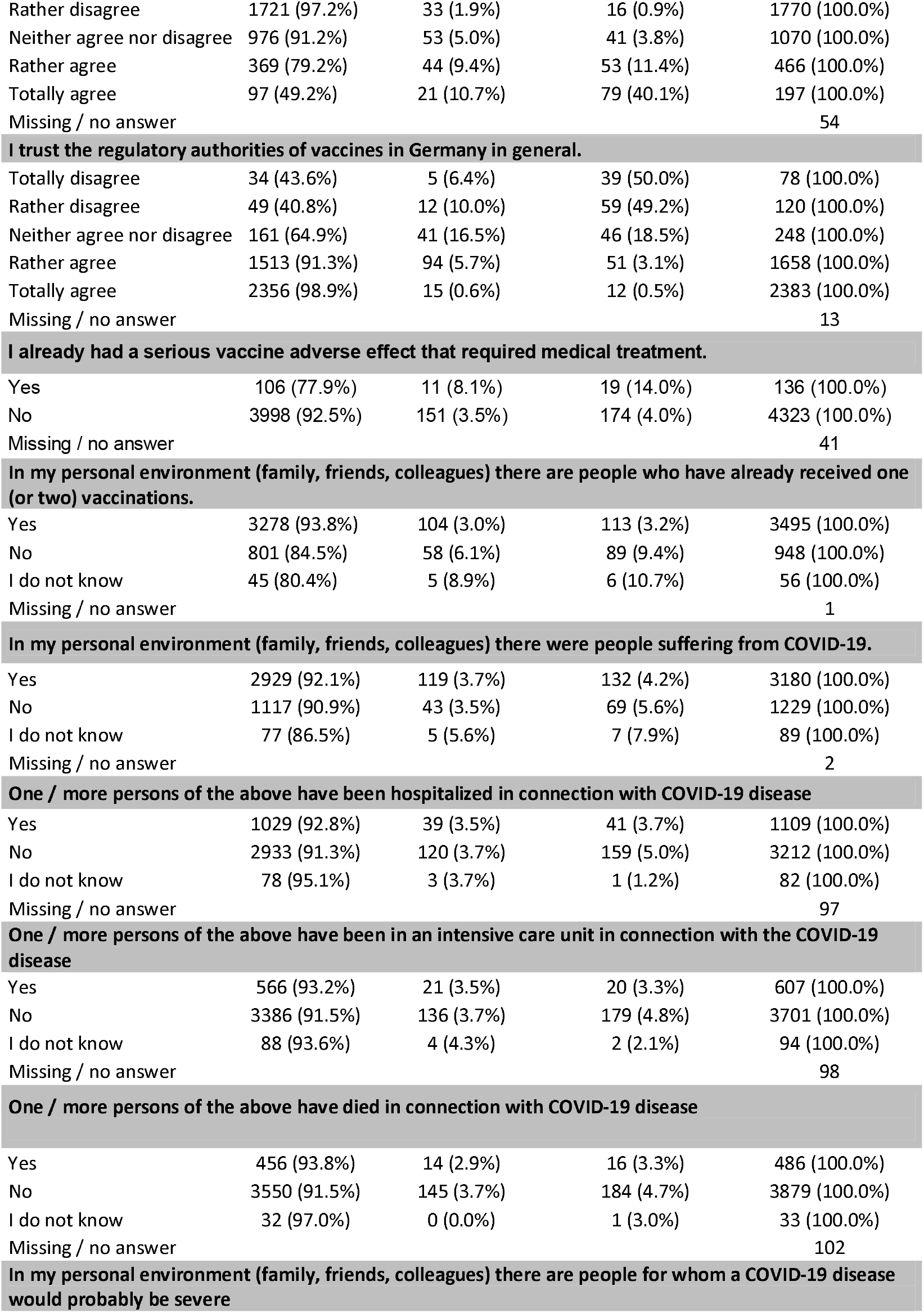
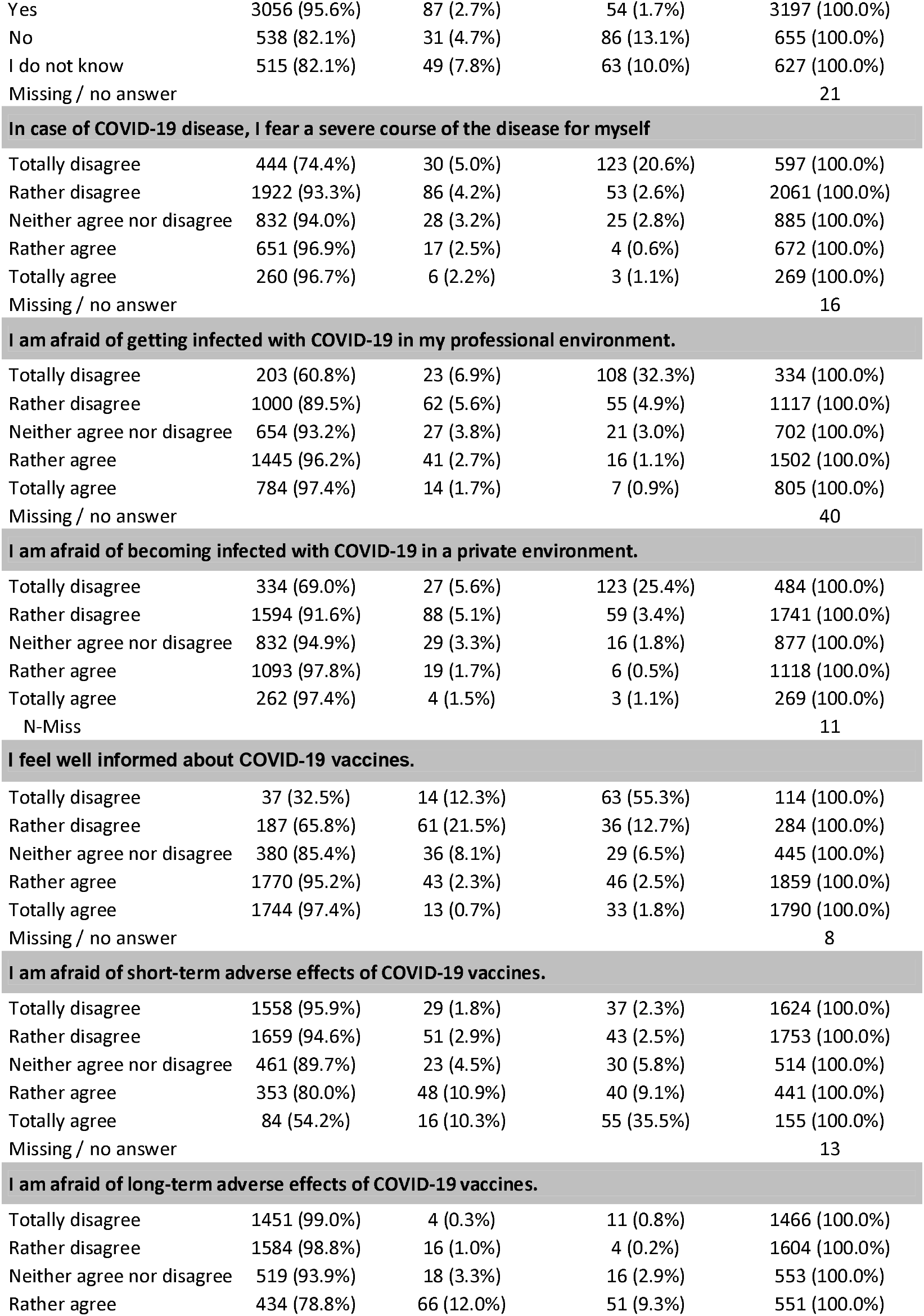
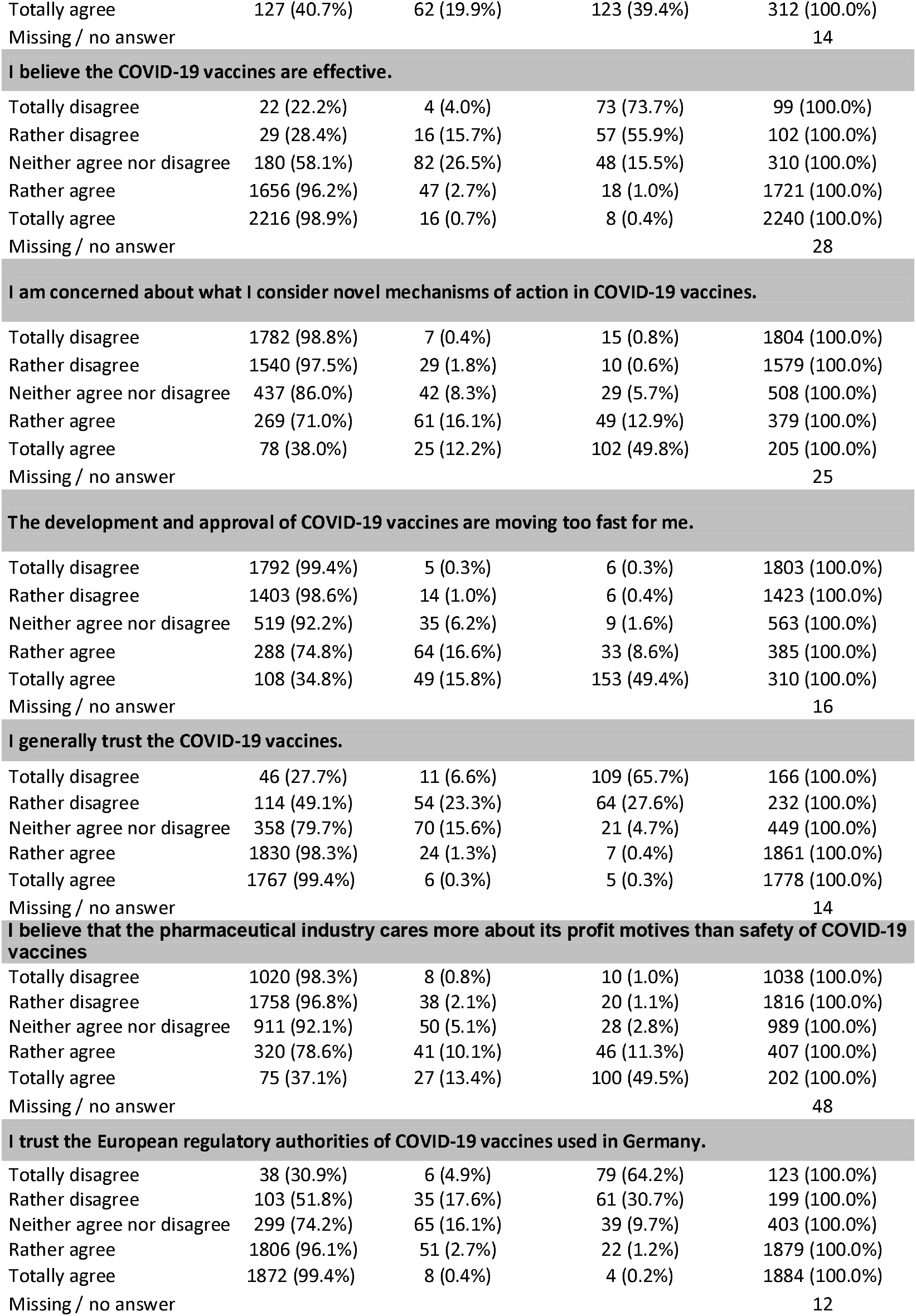
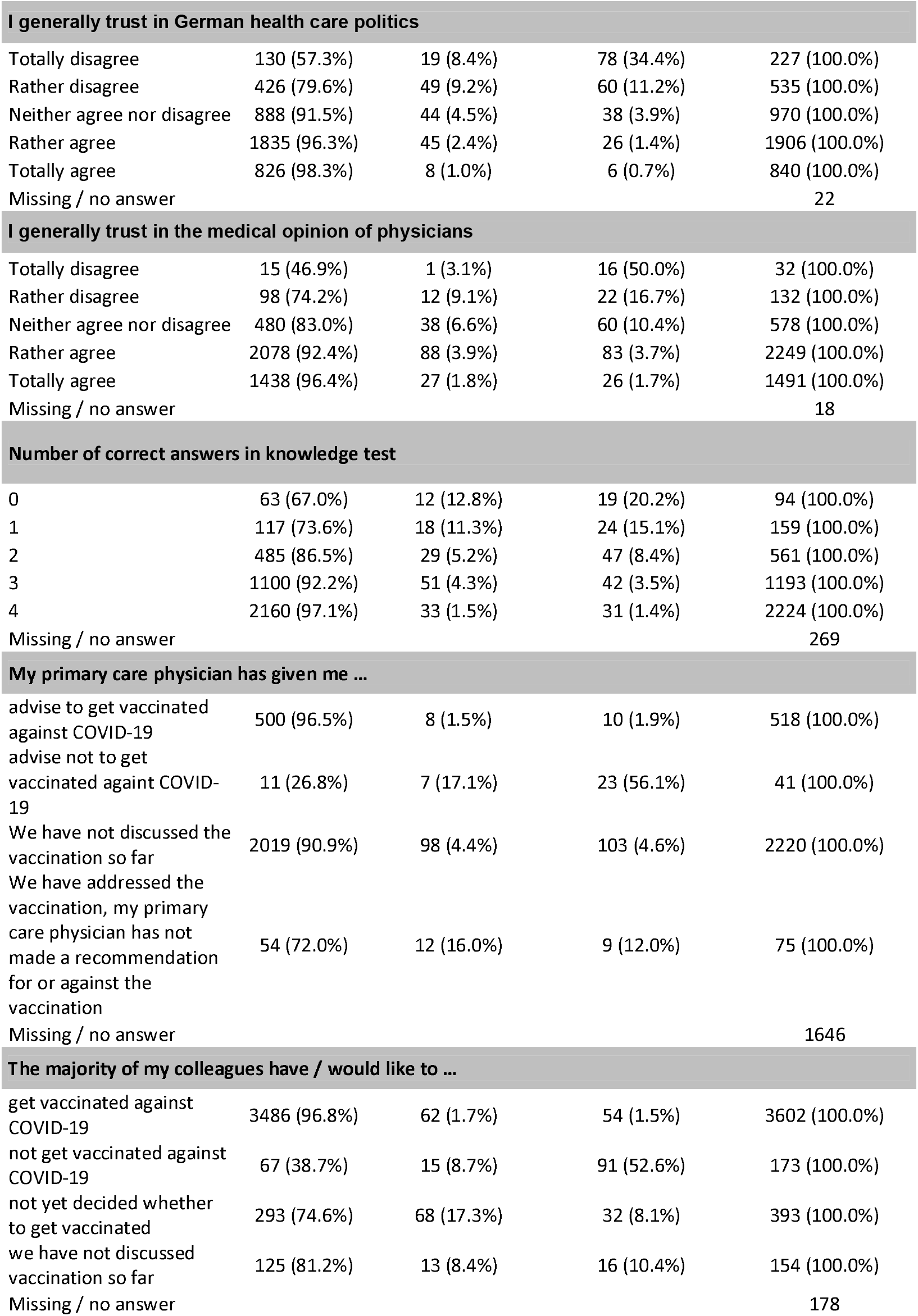
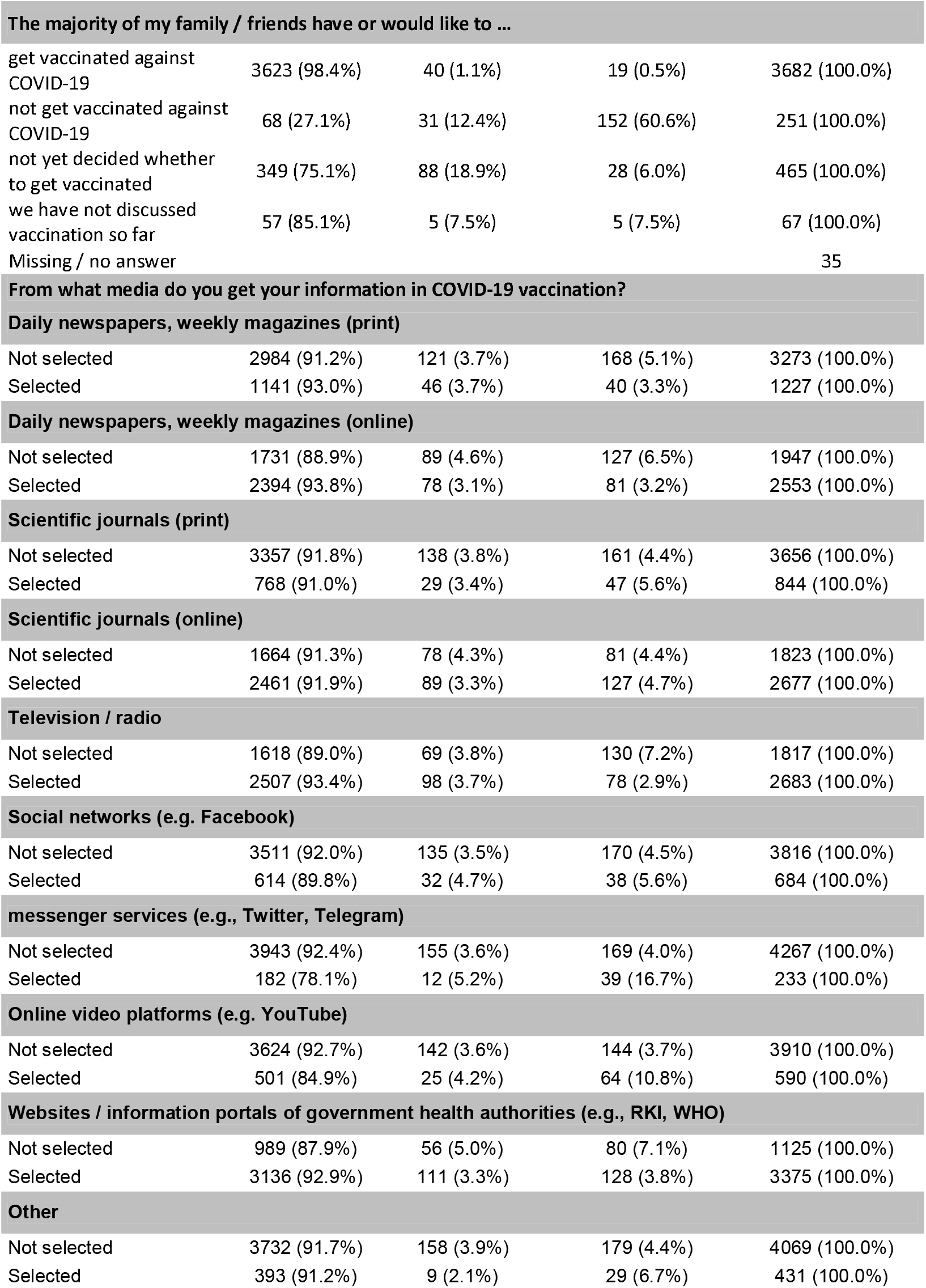
overall results.

|  | Want to get vaccinated or already vaccinated (N=4125) | Undecided (N=167) | Do not want to get vaccinated (N=208) | Total (N=4500) |
| --- | --- | --- | --- | --- |
| **Sex** |  |  |  |  |
| Female | 2392 (91.6%) | 115 (4.4%) | 103 (3.9%) | 2610 (100.0%) |
| Male | 1726 (91.9%) | 51 (2.7%) | 102 (5.4%) | 1879 (100.0%) |
| Missing / no answer |  |  |  | 11 |
| **Age** |  |  |  |  |
| ≤20 years | 134 (85.9%) | 8 (5.1%) | 14 (9.0%) | 156 (100.0%) |
| 21-30 years | 1491 (93.0%) | 51 (3.2%) | 61 (3.8%) | 1603 (100.0%) |
| 31-40 years | 794 (91.1%) | 39 (4.5%) | 39 (4.5%) | 872 (100.0%) |
| 41-50 years | 664 (91.0%) | 30 (4.1%) | 36 (4.9%) | 730 (100.0%) |
| 51-60 years | 714 (92.0%) | 29 (3.7%) | 33 (4.3%) | 776 (100.0%) |
| ≥61 years | 325 (91.5%) | 8 (2.3%) | 22 (6.2%) | 355 (100.0%) |
| Missing / no answer |  |  |  | 8 |

| Profession |  |  |  |  |
| --- | --- | --- | --- | --- |
| Certified nurse | 424 (91.0%) | 18 (3.9%) | 24 (5.2%) | 466 (100.0%) |
| Other non-physician medical staff | 311 (89.9%) | 17 (4.9%) | 18 (5.2%) | 346 (100.0%) |
| Resident | 302 (93.5%) | 9 (2.8%) | 12 (3.7%) | 323 (100.0%) |
| Physician with specialist / personnel responsibility | 1225 (92.2%) | 42 (3.2%) | 62 (4.7%) | 1329 (100.0%) |
| Administration / science | 241 (90.6%) | 16 (6.0%) | 9 (3.4%) | 266 (100.0%) |
| Medical student | 1225 (93.3%) | 37 (2.8%) | 51 (3.9%) |  |
| Dentist / dentistry student / dental assisting personnel | 241 (83.7%) | 21 (7.3%) | 26 (9.0%) | 288 (100.0%) |
| Other non-physician medical staff | 143 (96.6%) | 3 (2.0%) | 2 (1.4%) | 148 (100.0%) |
| Missing / no answer |  |  |  | 21 |
| Work setting |  |  |  |  |
| Maximum-care hospital / university hospital | 1807 (94.4%) | 55 (2.9%) | 53 (2.8%) | 1915 (100.0%) |
| Hospital of other care levels | 758 (93.8%) | 23 (2.8%) | 27 (3.3%) | 808 (100.0%) |
| Medical practice / medical care center | 781 (86.0%) | 49 (5.4%) | 78 (8.6%) | 908 (100.0%) |
| Rescue service | 126 (96.2%) | 3 (2.3%) | 2 (1.5%) | 131 (100.0%) |
| Nursing home / retirement home | 27 (100.0%) | 0 (0.0%) | 0 (0.0%) | 27 (100.0%) |
| Outpatient nursing service | 35 (77.8%) | 3 (6.7%) | 7 (15.6%) |  |
| Other non-physician medical staff | 263 (93.6%) | 10 (3.6%) | 8 (2.8%) | 281 (100.0%) |
| Missing / no answer |  |  |  | 385 |
| You indicated that you work in an inpatient setting. Do you work in an intensive care unit in this setting? |  |  |  |  |
| Yes | 1053 (95.3%) | 20 (1.8%) | 32 (2.9%) | 1105 (100.0%) |
| No | 1428 (93.6%) | 55 (3.6%) | 42 (2.8%) | 1525 (100.0%) |
| Missing / no answer |  |  |  | 1870 |
| I work with COVID-19 positive patients |  |  |  |  |
| Never | 1559 (90.2%) | 80 (4.6%) | 90 (5.2%) | 1729 (100.0%) |
| < 50% of working days | 1703 (93.8%) | 46 (2.5%) | 66 (3.6%) | 1815 (100.0%) |
| > 50% of working days (but not every working day) | 287 (93.5%) | 7 (2.3%) | 13 (4.2%) | 307 (100.0%) |
| On each working day | 161 (94.2%) | 2 (1.2%) | 8 (4.7%) | 171 (100.0%) |
| Missing / no answer |  |  |  | 478 |
| Region |  |  |  |  |
| Southern Germany | 1475 (93.2%) | 52 (3.3%) | 55 (3.5%) | 1582 (100.0%) |
| Eastern Germany | 344 (90.5%) | 15 (3.9%) | 21 (5.5%) | 380 (100.0%) |
| Northern Germany | 1132 (89.4%) | 57 (4.5%) | 77 (6.1%) | 1266 (100.0%) |
| Western Germany | 1083 (92.7%) | 37 (3.2%) | 48 (4.1%) | 1168 (100.0%) |
| Missing / no answer |  |  |  | 104 |
| **I make sure to keep my vaccinations up to date.** |  |  |  |  |
| Totally disagree | 14 (29.2%) | 4 (8.3%) | 30 (62.5%) | 48 (100.0%) |
| Rather disagree | 113 (73.9%) | 8 (5.2%) | 32 (20.9%) | 153 (100.0%) |
| Neither agree nor disagree | 107 (81.1%) | 11 (8.3%) | 14 (10.6%) | 132 (100.0%) |
| Rather agree | 1043 (89.4%) | 69 (5.9%) | 55 (4.7%) | 1167 (100.0%) |
| Totally agree | 2838 (95.1%) | 73 (2.4%) | 72 (2.4%) | 2983 (100.0%) |
| Missing / no answer |  |  |  | 17 |
| **I receive flu (influenza) vaccinations regularly** |  |  |  |  |
| Totally disagree | 722 (75.8%) | 85 (8.9%) | 146 (15.3%) | 953 (100.0%) |
| Rather disagree | 466 (91.4%) | 31 (6.1%) | 13 (2.5%) | 510 (100.0%) |
| Neither agree nor disagree | 271 (92.5%) | 9 (3.1%) | 13 (4.4%) | 293 (100.0%) |
| Rather agree | 758 (95.9%) | 19 (2.4%) | 13 (1.6%) | 790 (100.0%) |
| Totally agree | 1878 (97.7%) | 23 (1.2%) | 22 (1.1%) | 1923 (100.0%) |
| Missing / no answer |  |  |  | 31 |
| **I feel well informed about vaccines in general.** |  |  |  |  |
| Totally disagree | 30 (52.6%) | 8 (14.0%) | 19 (33.3%) | 57 (100.0%) |
| Rather disagree | 150 (72.1%) | 24 (11.5%) | 34 (16.3%) | 208 (100.0%) |
| Neither agree nor disagree | 331 (82.3%) | 43 (10.7%) | 28 (7.0%) | 402 (100.0%) |
| Rather agree | 1576 (92.9%) | 64 (3.8%) | 56 (3.3%) | 1696 (100.0%) |
| Totally agree | 2029 (95.3%) | 28 (1.3%) | 71 (3.3%) | 2128 (100.0%) |
| Missing / no answer |  |  |  | 9 |
| **I am generally afraid of adverse effects of vaccinations.** |  |  |  |  |
| Totally disagree | 1851 (96.3%) | 28 (1.5%) | 43 (2.2%) | 1922 (100.0%) |
| Rather disagree | 1678 (94.1%) | 56 (3.1%) | 49 (2.7%) | 1783 (100.0%) |
| Neither agree nor disagree | 254 (79.9%) | 29 (9.1%) | 35 (11.0%) | 318 (100.0%) |
| Rather agree | 241 (76.5%) | 39 (12.4%) | 35 (11.1%) | 315 (100.0%) |
| Totally agree | 95 (61.7%) | 15 (9.7%) | 44 (28.6%) | 154 (100.0%) |
| Missing / no answer |  |  |  | 8 |
| **I think vaccinations are generally effective.** |  |  |  |  |
| Totally disagree | 30 (66.7%) | 1 (2.2%) | 14 (31.1%) | 45 (100.0%) |
| Rather disagree | 13 (28.3%) | 5 (10.9%) | 28 (60.9%) | 46 (100.0%) |
| Neither agree nor disagree | 29 (39.2%) | 8 (10.8%) | 37 (50.0%) | 74 (100.0%) |
| Rather agree | 456 (79.6%) | 66 (11.5%) | 51 (8.9%) | 573 (100.0%) |
| Totally agree | 3594 (95.7%) | 87 (2.3%) | 76 (2.0%) | 3757 (100.0%) |
| Missing / no answer |  |  |  | 5 |
| **I trust vaccines in general.** |  |  |  |  |
| Totally disagree | 18 (31.0%) | 0 (0.0%) | 40 (69.0%) | 58 (100.0%) |
| Rather disagree | 30 (37.0%) | 15 (18.5%) | 36 (44.4%) | 81 (100.0%) |
| Neither agree nor disagree | 97 (62.2%) | 27 (17.3%) | 32 (20.5%) | 156 (100.0%) |
| Rather agree | 1038 (88.2%) | 80 (6.8%) | 59 (5.0%) | 1177 (100.0%) |
| Totally agree | 2937 (97.2%) | 45 (1.5%) | 40 (1.3%) | 3022 (100.0%) |
| Missing / no answer |  |  |  | 6 |
| **I believe that the pharmaceutical industry puts profit motives over safety of vaccines.** |  |  |  |  |
| Totally disagree | 914 (96.9%) | 12 (1.3%) | 17 (1.8%) | 943 (100.0%) |
| Rather disagree | 1721 (97.2%) | 33 (1.9%) | 16 (0.9%) | 1770 (100.0%) |
| Neither agree nor disagree | 976 (91.2%) | 53 (5.0%) | 41 (3.8%) | 1070 (100.0%) |
| Rather agree | 369 (79.2%) | 44 (9.4%) | 53 (11.4%) | 466 (100.0%) |
| Totally agree | 97 (49.2%) | 21 (10.7%) | 79 (40.1%) | 197 (100.0%) |
| Missing / no answer |  |  |  | 54 |
| **I trust the regulatory authorities of vaccines in Germany in general.** |  |  |  |  |
| Totally disagree | 34 (43.6%) | 5 (6.4%) | 39 (50.0%) | 78 (100.0%) |
| Rather disagree | 49 (40.8%) | 12 (10.0%) | 59 (49.2%) | 120 (100.0%) |
| Neither agree nor disagree | 161 (64.9%) | 41 (16.5%) | 46 (18.5%) | 248 (100.0%) |
| Rather agree | 1513 (91.3%) | 94 (5.7%) | 51 (3.1%) | 1658 (100.0%) |
| Totally agree | 2356 (98.9%) | 15 (0.6%) | 12 (0.5%) | 2383 (100.0%) |
| Missing / no answer |  |  |  | 13 |
| **I already had a serious vaccine adverse effect that required medical treatment.** |  |  |  |  |
| Yes | 106 (77.9%) | 11 (8.1%) | 19 (14.0%) | 136 (100.0%) |
| No | 3998 (92.5%) | 151 (3.5%) | 174 (4.0%) | 4323 (100.0%) |
| Missing / no answer |  |  |  | 41 |
| Yes | 3278 (93.8%) | 104 (3.0%) | 113 (3.2%) | 3495 (100.0%) |
| No | 801 (84.5%) | 58 (6.1%) | 89 (9.4%) | 948 (100.0%) |
| I do not know | 45 (80.4%) | 5 (8.9%) | 6 (10.7%) | 56 (100.0%) |
| Missing / no answer |  |  |  | 1 |
| **In my personal environment (family, friends, colleagues) there were people suffering from COVID-19.** |  |  |  |  |
| Yes | 2929 (92.1%) | 119 (3.7%) | 132 (4.2%) | 3180 (100.0%) |
| No | 1117 (90.9%) | 43 (3.5%) | 69 (5.6%) | 1229 (100.0%) |
| I do not know | 77 (86.5%) | 5 (5.6%) | 7 (7.9%) | 89 (100.0%) |
| Missing / no answer |  |  |  | 2 |
| **One / more persons of the above have been hospitalized in connection with COVID-19 disease** |  |  |  |  |
| Yes | 1029 (92.8%) | 39 (3.5%) | 41 (3.7%) | 1109 (100.0%) |
| No | 2933 (91.3%) | 120 (3.7%) | 159 (5.0%) | 3212 (100.0%) |
| I do not know | 78 (95.1%) | 3 (3.7%) | 1 (1.2%) | 82 (100.0%) |
| Missing / no answer |  |  |  | 97 |
| **One / more persons of the above have been in an intensive care unit in connection with the COVID-19 disease** |  |  |  |  |
| Yes | 566 (93.2%) | 21 (3.5%) | 20 (3.3%) | 607 (100.0%) |
| No | 3386 (91.5%) | 136 (3.7%) | 179 (4.8%) | 3701 (100.0%) |
| I do not know | 88 (93.6%) | 4 (4.3%) | 2 (2.1%) | 94 (100.0%) |
| Missing / no answer |  |  |  | 98 |
| **One / more persons of the above have died in connection with COVID-19 disease** |  |  |  |  |
| Yes | 456 (93.8%) | 14 (2.9%) | 16 (3.3%) | 486 (100.0%) |
| No | 3550 (91.5%) | 145 (3.7%) | 184 (4.7%) | 3879 (100.0%) |
| I do not know | 32 (97.0%) | 0 (0.0%) | 1 (3.0%) | 33 (100.0%) |
| Missing / no answer |  |  |  | 102 |
| Yes | 3056 (95.6%) | 87 (2.7%) | 54 (1.7%) | 3197 (100.0%) |
| No | 538 (82.1%) | 31 (4.7%) | 86 (13.1%) | 655 (100.0%) |
| I do not know | 515 (82.1%) | 49 (7.8%) | 63 (10.0%) | 627 (100.0%) |
| Missing / no answer |  |  |  | 21 |
| **In case of COVID-19 disease, I fear a severe course of the disease for myself** |  |  |  |  |
| Totally disagree | 444 (74.4%) | 30 (5.0%) | 123 (20.6%) | 597 (100.0%) |
| Rather disagree | 1922 (93.3%) | 86 (4.2%) | 53 (2.6%) | 2061 (100.0%) |
| Neither agree nor disagree | 832 (94.0%) | 28 (3.2%) | 25 (2.8%) | 885 (100.0%) |
| Rather agree | 651 (96.9%) | 17 (2.5%) | 4 (0.6%) | 672 (100.0%) |
| Totally agree | 260 (96.7%) | 6 (2.2%) | 3 (1.1%) | 269 (100.0%) |
| Missing / no answer |  |  |  | 16 |
| **I am afraid of getting infected with COVID-19 in my professional environment.** |  |  |  |  |
| Totally disagree | 203 (60.8%) | 23 (6.9%) | 108 (32.3%) | 334 (100.0%) |
| Rather disagree | 1000 (89.5%) | 62 (5.6%) | 55 (4.9%) | 1117 (100.0%) |
| Neither agree nor disagree | 654 (93.2%) | 27 (3.8%) | 21 (3.0%) | 702 (100.0%) |
| Rather agree | 1445 (96.2%) | 41 (2.7%) | 16 (1.1%) | 1502 (100.0%) |
| Totally agree | 784 (97.4%) | 14 (1.7%) | 7 (0.9%) | 805 (100.0%) |
| Missing / no answer |  |  |  | 40 |
| **I am afraid of becoming infected with COVID-19 in a private environment.** |  |  |  |  |
| Totally disagree | 334 (69.0%) | 27 (5.6%) | 123 (25.4%) | 484 (100.0%) |
| Rather disagree | 1594 (91.6%) | 88 (5.1%) | 59 (3.4%) | 1741 (100.0%) |
| Neither agree nor disagree | 832 (94.9%) | 29 (3.3%) | 16 (1.8%) | 877 (100.0%) |
| Rather agree | 1093 (97.8%) | 19 (1.7%) | 6 (0.5%) | 1118 (100.0%) |
| Totally agree | 262 (97.4%) | 4 (1.5%) | 3 (1.1%) | 269 (100.0%) |
| N-Miss |  |  |  | 11 |
| **I feel well informed about COVID-19 vaccines.** |  |  |  |  |
| Totally disagree | 37 (32.5%) | 14 (12.3%) | 63 (55.3%) | 114 (100.0%) |
| Rather disagree | 187 (65.8%) | 61 (21.5%) | 36 (12.7%) | 284 (100.0%) |
| Neither agree nor disagree | 380 (85.4%) | 36 (8.1%) | 29 (6.5%) | 445 (100.0%) |
| Rather agree | 1770 (95.2%) | 43 (2.3%) | 46 (2.5%) | 1859 (100.0%) |
| Totally agree | 1744 (97.4%) | 13 (0.7%) | 33 (1.8%) | 1790 (100.0%) |
| Missing / no answer |  |  |  | 8 |
| **I am afraid of short-term adverse effects of COVID-19 vaccines.** |  |  |  |  |
| Totally disagree | 1558 (95.9%) | 29 (1.8%) | 37 (2.3%) | 1624 (100.0%) |
| Rather disagree | 1659 (94.6%) | 51 (2.9%) | 43 (2.5%) | 1753 (100.0%) |
| Neither agree nor disagree | 461 (89.7%) | 23 (4.5%) | 30 (5.8%) | 514 (100.0%) |
| Rather agree | 353 (80.0%) | 48 (10.9%) | 40 (9.1%) | 441 (100.0%) |
| Totally agree | 84 (54.2%) | 16 (10.3%) | 55 (35.5%) | 155 (100.0%) |
| Missing / no answer |  |  |  | 13 |
| **I am afraid of long-term adverse effects of COVID-19 vaccines.** |  |  |  |  |
| Totally disagree | 1451 (99.0%) | 4 (0.3%) | 11 (0.8%) | 1466 (100.0%) |
| Rather disagree | 1584 (98.8%) | 16 (1.0%) | 4 (0.2%) | 1604 (100.0%) |
| Neither agree nor disagree | 519 (93.9%) | 18 (3.3%) | 16 (2.9%) | 553 (100.0%) |
| Rather agree | 434 (78.8%) | 66 (12.0%) | 51 (9.3%) | 551 (100.0%) |
| Totally agree | 127 (40.7%) | 62 (19.9%) | 123 (39.4%) | 312 (100.0%) |
| Missing / no answer |  |  |  | 14 |
| **I believe the COVID-19 vaccines are effective.** |  |  |  |  |
| Totally disagree | 22 (22.2%) | 4 (4.0%) | 73 (73.7%) | 99 (100.0%) |
| Rather disagree | 29 (28.4%) | 16 (15.7%) | 57 (55.9%) | 102 (100.0%) |
| Neither agree nor disagree | 180 (58.1%) | 82 (26.5%) | 48 (15.5%) | 310 (100.0%) |
| Rather agree | 1656 (96.2%) | 47 (2.7%) | 18 (1.0%) | 1721 (100.0%) |
| Totally agree | 2216 (98.9%) | 16 (0.7%) | 8 (0.4%) | 2240 (100.0%) |
| Missing / no answer |  |  |  | 28 |
| **I am concerned about what I consider novel mechanisms of action in COVID-19 vaccines.** |  |  |  |  |
| Totally disagree | 1782 (98.8%) | 7 (0.4%) | 15 (0.8%) | 1804 (100.0%) |
| Rather disagree | 1540 (97.5%) | 29 (1.8%) | 10 (0.6%) | 1579 (100.0%) |
| Neither agree nor disagree | 437 (86.0%) | 42 (8.3%) | 29 (5.7%) | 508 (100.0%) |
| Rather agree | 269 (71.0%) | 61 (16.1%) | 49 (12.9%) | 379 (100.0%) |
| Totally agree | 78 (38.0%) | 25 (12.2%) | 102 (49.8%) | 205 (100.0%) |
| Missing / no answer |  |  |  | 25 |
| **The development and approval of COVID-19 vaccines are moving too fast for me.** |  |  |  |  |
| Totally disagree | 1792 (99.4%) | 5 (0.3%) | 6 (0.3%) | 1803 (100.0%) |
| Rather disagree | 1403 (98.6%) | 14 (1.0%) | 6 (0.4%) | 1423 (100.0%) |
| Neither agree nor disagree | 519 (92.2%) | 35 (6.2%) | 9 (1.6%) | 563 (100.0%) |
| Rather agree | 288 (74.8%) | 64 (16.6%) | 33 (8.6%) | 385 (100.0%) |
| Totally agree | 108 (34.8%) | 49 (15.8%) | 153 (49.4%) | 310 (100.0%) |
| Missing / no answer |  |  |  | 16 |
| **I generally trust the COVID-19 vaccines.** |  |  |  |  |
| Totally disagree | 46 (27.7%) | 11 (6.6%) | 109 (65.7%) | 166 (100.0%) |
| Rather disagree | 114 (49.1%) | 54 (23.3%) | 64 (27.6%) | 232 (100.0%) |
| Neither agree nor disagree | 358 (79.7%) | 70 (15.6%) | 21 (4.7%) | 449 (100.0%) |
| Rather agree | 1830 (98.3%) | 24 (1.3%) | 7 (0.4%) | 1861 (100.0%) |
| Totally agree | 1767 (99.4%) | 6 (0.3%) | 5 (0.3%) | 1778 (100.0%) |
| Missing / no answer |  |  |  | 14 |
| **I believe that the pharmaceutical industry cares more about its profit motives than safety of COVID-19 vaccines** |  |  |  |  |
| Totally disagree | 1020 (98.3%) | 8 (0.8%) | 10 (1.0%) | 1038 (100.0%) |
| Rather disagree | 1758 (96.8%) | 38 (2.1%) | 20 (1.1%) | 1816 (100.0%) |
| Neither agree nor disagree | 911 (92.1%) | 50 (5.1%) | 28 (2.8%) | 989 (100.0%) |
| Rather agree | 320 (78.6%) | 41 (10.1%) | 46 (11.3%) | 407 (100.0%) |
| Totally agree | 75 (37.1%) | 27 (13.4%) | 100 (49.5%) | 202 (100.0%) |
| Missing / no answer |  |  |  | 48 |
| **I trust the European regulatory authorities of COVID-19 vaccines used in Germany.** |  |  |  |  |
| Totally disagree | 38 (30.9%) | 6 (4.9%) | 79 (64.2%) | 123 (100.0%) |
| Rather disagree | 103 (51.8%) | 35 (17.6%) | 61 (30.7%) | 199 (100.0%) |
| Neither agree nor disagree | 299 (74.2%) | 65 (16.1%) | 39 (9.7%) | 403 (100.0%) |
| Rather agree | 1806 (96.1%) | 51 (2.7%) | 22 (1.2%) | 1879 (100.0%) |
| Totally agree | 1872 (99.4%) | 8 (0.4%) | 4 (0.2%) | 1884 (100.0%) |
| Missing / no answer |  |  |  | 12 |

| **I generally trust in German health care politics** |  |  |  |  |
| --- | --- | --- | --- | --- |
| Totally disagree | 130 (57.3%) | 19 (8.4%) | 78 (34.4%) | 227 (100.0%) |
| Rather disagree | 426 (79.6%) | 49 (9.2%) | 60 (11.2%) | 535 (100.0%) |
| Neither agree nor disagree | 888 (91.5%) | 44 (4.5%) | 38 (3.9%) | 970 (100.0%) |
| Rather agree | 1835 (96.3%) | 45 (2.4%) | 26 (1.4%) | 1906 (100.0%) |
| Totally agree | 826 (98.3%) | 8 (1.0%) | 6 (0.7%) | 840 (100.0%) |
| Missing / no answer |  |  |  | 22 |
| **I generally trust in the medical opinion of physicians** |  |  |  |  |
| Totally disagree | 15 (46.9%) | 1 (3.1%) | 16 (50.0%) | 32 (100.0%) |
| Rather disagree | 98 (74.2%) | 12 (9.1%) | 22 (16.7%) | 132 (100.0%) |
| Neither agree nor disagree | 480 (83.0%) | 38 (6.6%) | 60 (10.4%) | 578 (100.0%) |
| Rather agree | 2078 (92.4%) | 88 (3.9%) | 83 (3.7%) | 2249 (100.0%) |
| Totally agree | 1438 (96.4%) | 27 (1.8%) | 26 (1.7%) | 1491 (100.0%) |
| Missing / no answer |  |  |  | 18 |
| **Number of correct answers in knowledge test** |  |  |  |  |
| 0 | 63 (67.0%) | 12 (12.8%) | 19 (20.2%) | 94 (100.0%) |
| 1 | 117 (73.6%) | 18 (11.3%) | 24 (15.1%) | 159 (100.0%) |
| 2 | 485 (86.5%) | 29 (5.2%) | 47 (8.4%) | 561 (100.0%) |
| 3 | 1100 (92.2%) | 51 (4.3%) | 42 (3.5%) | 1193 (100.0%) |
| 4 | 2160 (97.1%) | 33 (1.5%) | 31 (1.4%) | 2224 (100.0%) |
| Missing / no answer |  |  |  | 269 |
| **My primary care physician has given me ...** |  |  |  |  |
| advise to get vaccinated against COVID-19 | 500 (96.5%) | 8 (1.5%) | 10 (1.9%) | 518 (100.0%) |
| advise not to get vaccinated against COVID-19 | 11 (26.8%) | 7 (17.1%) | 23 (56.1%) | 41 (100.0%) |
| We have not discussed the vaccination so far | 2019 (90.9%) | 98 (4.4%) | 103 (4.6%) | 2220 (100.0%) |
| We have addressed the vaccination, my primary care physician has not made a recommendation for or against the vaccination | 54 (72.0%) | 12 (16.0%) | 9 (12.0%) | 75 (100.0%) |
| Missing / no answer |  |  |  | 1646 |
| **The majority of my colleagues have / would like to ...** |  |  |  |  |
| get vaccinated against COVID-19 | 3486 (96.8%) | 62 (1.7%) | 54 (1.5%) | 3602 (100.0%) |
| not get vaccinated against COVID-19 | 67 (38.7%) | 15 (8.7%) | 91 (52.6%) | 173 (100.0%) |
| not yet decided whether to get vaccinated | 293 (74.6%) | 68 (17.3%) | 32 (8.1%) | 393 (100.0%) |
| we have not discussed vaccination so far | 125 (81.2%) | 13 (8.4%) | 16 (10.4%) | 154 (100.0%) |
| Missing / no answer |  |  |  | 178 |

| The majority of my family / friends have or would like to ... |  |  |  |  |
| --- | --- | --- | --- | --- |
| get vaccinated against COVID-19 | 3623 (98.4%) | 40 (1.1%) | 19 (0.5%) | 3682 (100.0%) |
| not get vaccinated against COVID-19 | 68 (27.1%) | 31 (12.4%) | 152 (60.6%) | 251 (100.0%) |
| not yet decided whether to get vaccinated | 349 (75.1%) | 88 (18.9%) | 28 (6.0%) | 465 (100.0%) |
| we have not discussed vaccination so far | 57 (85.1%) | 5 (7.5%) | 5 (7.5%) | 67 (100.0%) |
| Missing / no answer |  |  |  | 35 |
| From what media do you get your information in COVID-19 vaccination? |  |  |  |  |
| Daily newspapers, weekly magazines (print) |  |  |  |  |
| Not selected | 2984 (91.2%) | 121 (3.7%) | 168 (5.1%) | 3273 (100.0%) |
| Selected | 1141 (93.0%) | 46 (3.7%) | 40 (3.3%) | 1227 (100.0%) |
| Daily newspapers, weekly magazines (online) |  |  |  |  |
| Not selected | 1731 (88.9%) | 89 (4.6%) | 127 (6.5%) | 1947 (100.0%) |
| Selected | 2394 (93.8%) | 78 (3.1%) | 81 (3.2%) | 2553 (100.0%) |
| Scientific journals (print) |  |  |  |  |
| Not selected | 3357 (91.8%) | 138 (3.8%) | 161 (4.4%) | 3656 (100.0%) |
| Selected | 768 (91.0%) | 29 (3.4%) | 47 (5.6%) | 844 (100.0%) |
| Scientific journals (online) |  |  |  |  |
| Not selected | 1664 (91.3%) | 78 (4.3%) | 81 (4.4%) | 1823 (100.0%) |
| Selected | 2461 (91.9%) | 89 (3.3%) | 127 (4.7%) | 2677 (100.0%) |
| Television / radio |  |  |  |  |
| Not selected | 1618 (89.0%) | 69 (3.8%) | 130 (7.2%) | 1817 (100.0%) |
| Selected | 2507 (93.4%) | 98 (3.7%) | 78 (2.9%) | 2683 (100.0%) |
| Social networks (e.g. Facebook) |  |  |  |  |
| Not selected | 3511 (92.0%) | 135 (3.5%) | 170 (4.5%) | 3816 (100.0%) |
| Selected | 614 (89.8%) | 32 (4.7%) | 38 (5.6%) | 684 (100.0%) |
| messenger services (e.g., Twitter, Telegram) |  |  |  |  |
| Not selected | 3943 (92.4%) | 155 (3.6%) | 169 (4.0%) | 4267 (100.0%) |
| Selected | 182 (78.1%) | 12 (5.2%) | 39 (16.7%) | 233 (100.0%) |
| Online video platforms (e.g. YouTube) |  |  |  |  |
| Not selected | 3624 (92.7%) | 142 (3.6%) | 144 (3.7%) | 3910 (100.0%) |
| Selected | 501 (84.9%) | 25 (4.2%) | 64 (10.8%) | 590 (100.0%) |
| Websites / information portals of government health authorities (e.g., RKI, WHO) |  |  |  |  |
| Not selected | 989 (87.9%) | 56 (5.0%) | 80 (7.1%) | 1125 (100.0%) |
| Selected | 3136 (92.9%) | 111 (3.3%) | 128 (3.8%) | 3375 (100.0%) |
| Other |  |  |  |  |
| Not selected | 3732 (91.7%) | 158 (3.9%) | 179 (4.4%) | 4069 (100.0%) |
| Selected | 393 (91.2%) | 9 (2.1%) | 29 (6.7%) | 431 (100.0%) |

**Supplemental Table 4:** Attitudes and experiences with vaccinations, Odds ratios with corresponding 95% confidence intervals. Large values indicate higher odds for willingness to vaccinate. If not stated otherwise “neither agree nor disagree” was used as reference.

| Predictor | Odds ratio | 95% confidence interval |  |
| --- | --- | --- | --- |
|  |  | Lower | Upper |
| I trust the regulatory authorities of vaccines in Germany in general. |  |  |  |
| Totally disagree | 0.42 | 0.25 | 0.70 |
| Rather disagree | 0.37 | 0.24 | 0.58 |
| Rather agree | 5.64 | 4.13 | 7.70 |
| Totally agree | 47.15 | 29.76 | 74.72 |
| I trust the European regulatory authorities of COVID-19 vaccines used in Germany. |  |  |  |
| Totally disagree | 0.16 | 0.10 | 0.24 |
| Rather disagree | 0.37 | 0.26 | 0.53 |
| Rather agree | 8.61 | 6.23 | 11.89 |
| Totally agree | 54.26 | 29.49 | 99.85 |
| I basically trust in the German health care politics. |  |  |  |
| Totally disagree | 0.12 | 0.09 | 0.18 |
| Rather disagree | 0.36 | 0.26 | 0.49 |
| Rather agree | 2.39 | 1.72 | 3.31 |
| Totally agree | 5.45 | 3.07 | 9.68 |
| I generally trust in the medical opinion of physicians. |  |  |  |
| Totally disagree | 0.18 | 0.09 | 0.37 |
| Rather disagree | 0.59 | 0.38 | 0.92 |
| Rather agree | 2.48 | 1.90 | 3.24 |
| Totally agree | 5.54 | 3.90 | 7.86 |
| I already had a serious vaccine adverse effect that required medical treatment. (Ref: No) |  |  |  |
| Yes | 0.29 | 0.19 | 0.44 |
| I make sure to keep my vaccinations up to date. |  |  |  |
| Totally disagree | 0.10 | 0.05 | 0.21 |
| Rather disagree | 0.66 | 0.38 | 1.16 |
| Rather agree | 1.97 | 1.22 | 3.16 |
| Totally agree | 4.57 | 2.87 | 7.29 |
| I receive flu (influenza) vaccinations regularly. |  |  |  |
| Totally disagree | 0.25 | 0.16 | 0.40 |
| Rather disagree | 0.86 | 0.50 | 1.47 |
| Rather agree | 1.92 | 1.10 | 3.37 |
| Totally agree | 3.39 | 2.00 | 5.73 |
| I feel well informed about COVID-19 vaccines. |  |  |  |
| Totally disagree | 0.08 | 0.05 | 0.13 |
| Rather disagree | 0.33 | 0.23 | 0.47 |
| Rather agree | 3.40 | 2.43 | 4.77 |
| Totally agree | 6.49 | 4.38 | 9.61 |
| I believe the COVID-19 vaccines are effective. |  |  |  |
| Totally disagree | 0.21 | 0.12 | 0.35 |
| Rather disagree | 0.29 | 0.18 | 0.47 |
| Rather agree | 18.40 | 13.16 | 25.73 |
| Totally agree | 66.69 | 42.05 | 105.76 |
| I generally trust COVID-19 vaccines. |  |  |  |
| Totally disagree | 0.10 | 0.06 | 0.15 |
| Rather disagree | 0.25 | 0.17 | 0.35 |
| Rather agree | 15.01 | 9.83 | 22.91 |
| Totally agree | 40.83 | 21.62 | 77.12 |
| I believe the pharmaceutical industry cares more about its profit motives than safety of COVID-19 vaccines. |  |  |  |
| Totally disagree | 4.85 | 2.88 | 8.16 |
| Rather disagree | 2.60 | 1.83 | 3.68 |
| Rather agree | 0.31 | 0.23 | 0.44 |
| Totally agree | 0.05 | 0.04 | 0.07 |
| I am afraid of short-term adverse effects of COVID-19 vaccines. |  |  |  |
| Totally disagree | 2.71 | 1.86 | 3.95 |
| Rather disagree | 2.03 | 1.43 | 2.89 |
| Rather agree | 0.46 | 0.32 | 0.67 |
| Totally agree | 0.14 | 0.09 | 0.21 |
| I am afraid of long-term adverse effects from COVID-19 vaccines. |  |  |  |
| Totally disagree | 6.34 | 3.42 | 11.73 |
| Rather disagree | 5.19 | 2.96 | 9.09 |
| Rather agree | 0.24 | 0.16 | 0.36 |
| Totally agree | 0.04 | 0.03 | 0.07 |
| I am concerned about what I consider novel mechanisms of action in COVID-19 vaccines. |  |  |  |
| Totally disagree | 13.16 | 8.07 | 21.47 |
| Rather disagree | 6.42 | 4.28 | 9.62 |
| Rather agree | 0.40 | 0.28 | 0.56 |
| Totally agree | 0.10 | 0.07 | 0.15 |
| The development and approval of COVID-19 vaccines are moving too fast for me. |  |  |  |
| Totally disagree | 13.81 | 7.08 | 26.93 |
| Rather disagree | 5.95 | 3.47 | 10.19 |
| Rather agree | 0.25 | 0.17 | 0.37 |
| Totally agree | 0.05 | 0.03 | 0.07 |
| I feel well informed about vaccines in general. |  |  |  |
| Totally disagree | 0.24 | 0.13 | 0.43 |
| Rather disagree | 0.55 | 0.37 | 0.83 |
| Rather agree | 2.82 | 2.05 | 3.87 |
| Totally agree | 4.40 | 3.17 | 6.09 |
| I think vaccinations are generally effective |  |  |  |
| Totally disagree | 3.10 | 1.43 | 6.74 |
| Rather disagree | 0.61 | 0.28 | 1.35 |
| Rather agree | 6.05 | 3.64 | 10.06 |
| Totally agree | 34.21 | 20.91 | 55.98 |
| I trust vaccines in general. |  |  |  |
| Totally disagree | 0.27 | 0.14 | 0.52 |
| Rather disagree | 0.36 | 0.21 | 0.62 |
| Rather agree | 4.54 | 3.14 | 6.57 |
| Totally agree | 21.02 | 14.25 | 31.01 |
| I am generally afraid of adverse effects of vaccinations. |  |  |  |
| Totally disagree | 6.57 | 4.57 | 9.44 |
| Rather disagree | 4.03 | 2.87 | 5.64 |
| Rather agree | 0.82 | 0.56 | 1.20 |
| Totally agree | 0.41 | 0.27 | 0.62 |
| I believe that the pharmaceutical industry's puts profit motives over the safety of vaccines. |  |  |  |
| Totally disagree | 3.04 | 1.98 | 4.65 |
| Rather disagree | 3.38 | 2.37 | 4.82 |
| Rather agree | 0.37 | 0.27 | 0.50 |
| Totally agree | 0.09 | 0.07 | 0.13 |

